# Pride and adversity among nurses and physicians during the pandemic in two US healthcare systems: a mixed methods analysis

**DOI:** 10.1101/2022.03.16.22272485

**Authors:** Igor Burstyn, Karyn Holt

## Abstract

Our aims were to examine themes of the most difficult or distressing events reported by healthcare workers during the first wave of COVID-19 pandemic in two US health care systems in order to identify common themes and to relate them to both behavioral theory and measures of anxiety and depression.

We conducted a cross-sectional survey during the early phases of the COIVD-19 pandemic in the US. We measured symptoms of anxiety and depression separately, captured demographics, and asked two open-ended questions regarding events that were the most difficult or stressful, and reinforced pride. The open-end questions were independently coded into themes developed by the authors and mapped to factors related to fostering well-being according to the Self-Determination Theory.

We recruited 874 nurses and 248 physicians. About a half shared their most distressing experiences as well as those experiences they were most proud of related to their professions. Themes that emerged from these narratives were congruent with prediction of Self-Determination theory that autonomy-supportive experiences will foster pride, while autonomy-thwarting experiences will cause distress. Those who reported distressful events were more anxious and depressed compared to those who did not. Among those who reported incidences that reinforced pride in the profession, depression was rarer compared to those who did not. These trends were evident after allowing for medical history and other covariates in logistic regressions.

Causal claims from our analysis should be made with caution due to the research design, a cross-sectional study design. Understanding of perceptions of the pandemic by nurses and physicians may help identify sources of distress and means of reinforcing pride in the professions, thereby helping nurses and physicians cope with disasters, and shape workplace policies during disasters that foster well-being among first responders.

No Patient or Public Contribution: We studied physicians and nurses themselves.

## Introduction

Evidence is robust that nurses and physicians around the world suffered from mental health distress, including anxiety and depression, during the COVID-19 pandemic (Pappa et al., 2020; Saragih et al., 2021; Wu et al., 2021). However, the specific perceptions of the events by nurses and physicians related to these mental health challenges are poorly captured, which may hinder effective interventions. Our prior analysis of risk factors for anxiety and depression among nurses and physicians during the first wave of COVID-19 in two health care systems in the US, who are also subjects in this report, posited that concern about contracting COVID-19 was a correlate of both anxiety and depression, especially among those who experienced recent bouts of poor health (Burstyn & Holt, 2021). Furthermore, the risk of anxiety and depression was reduced among those who felt competent using personal protective equipment and had access to it, reported few changes to working hours, and were surrounded by sufficient numbers of colleagues who were not seem as stressed. The expected support of immediate family and religious communities were protective. We did not delve into the specifics of experiences in the first report but only captured them on visual-analogue scales, thus potentially missing important features that contributed to mood disorders. We speculated that the “impact of work organization on anxiety and depression … may be related to the role of the safety climate or culture in moderating impact of the pandemic on work-induced mental health issues” (Dollard et al., 2019; Dollard & Bakker, 2010; Rickard et al., 2012), and that Self-Determination Theory (SDT) may offer a suitable framework for addressing these issues (Burstyn & Holt, 2021).

In this manuscript, thematic analysis of self-reported experiences of nurses and physicians during the first wave of COVID-19 is interpreted within the SDT framework (Ryan & Deci, 2017). According to the SDT, if changes are perceived as fostering a person’s sense of *autonomy* (e.g., having at least some latitude or input in the nature of changes), *competence* (e.g., leading to high level of professional performance), and *relatedness* (e.g., strengthening bond with colleagues and community at large), the people intrinsically cooperate with the change and perceive them as positive (experience wellness). On the other hand, if the psychological needs for autonomy, competence and relatedness are thwarted due to perceived coercive means employed to implement changes, then people become de-motivated and recognize changes as negative (experience ill-being). In the context of COVID-19 pandemic, there is evidence from longitudinal study that experience of autonomy-thwarting environment (i.e. frustration of basic psychological needs) among university students was the main predictor of depressive symptoms, after accounting for history of depressive symptoms (Levine et al., 2021). Need frustration was also were related to risk of mood disorders in a longitudinal analysis of adults experiencing COVID-19 lockdown in Belgium (Vermote et al., 2021). The COVID-19 pandemic precipitated changes in healthcare that necessitated changes in how nurses and physicians lived and practiced their professions. The extent to which such changes were positively received and the consequent ease of their acceptance and adoption, can be related to the perception of these changes as either autonomy-supportive or coercive. Although we did not formally access whether nurses and physicians experienced an autonomy-supportive vs. coercive workplace climate during implementation of changes resulting from COVID-19 pandemic, we can document whether their most difficult experiences (ill-being) correspond to their autonomy, competence, and relatedness of being threatened. Conversely, we can document whether their experiences that instilled or affirmed pride in their profession (wellness) corresponded to experiences that reinforced their autonomy, competence, and relatedness. Thus, although we are limited by not having employed psychometric scales typically used in evaluation of SDT (e.g., Work Climate and Problem at Work questionnaires), we have mapped themes identified in the narratives to psychological needs postulated by SDT.

We aimed to identify common themes among the most difficult or distressing events as well as experiences that instilled or reinforced a sense of pride reported by nurses and physicians and relate them to anxiety and depression during the first wave of COVID-19 pandemic in two US health care systems, one in the Northeast, the other in the West.

## Methods and materials

Our project received ethics approval from the Institutional Review Boards of the respective institutions (Drexel University and University of Nevada, Las Vegas).

## Study design and data collection

Details of the survey methodology and participating healthcare systems can be found in our earlier publication (Burstyn & Holt, 2021). We conducted a cross-sectional survey of all physicians and nurses employed and contracted by the Tower Health in Southeastern Pennsylvania (TH) and the University Medical Center, Las Vegas, Nevada (UMC), and licensed to practice in these states, corresponding to the early phases of the COVID-19 pandemic in the US. We recruited through Health Systems’ employee databases, distributing the invitation to enroll in the study and subsequent reminders by email with links to online surveys. Participation was voluntary, without reward for participation, and confidential. On June 3, 2020, we distributed invitations to TH survey to 203 advanced nurse practitioners and 4,336 registered nurses; at the same time, we distributed invitation to TH survey aimed at physicians to 2,496 active medical staff and 204 physician assistants. On September 9, 2020, we distributed invitations to the UMC version of the survey to 1,518 registered nurses and nurse practitioners, and 1,186 physicians.

We measured symptoms of anxiety and depression separately via a well-established the Hospital Anxiety and Depression Scale (HADS); scores of equal to or above 11 (range 0-21) indicate presence of these conditions (“case”) but are not clinical diagnoses (Bjelland et al., 2002; Zigmond & Snaith, 1983). We recorded age, marital status, gender, children under 18 years of age living at home, profession (nurse or physician), healthcare system, history of anxiety and depression prior to the pandemic (and evidence of exacerbation requiring treatment a year before the pandemic), report of positive COVID-19 test, whether respondent “have you had an episode when you have been unwell for two or more consecutive days” (whether or not they reported for duty) since the start of the pandemic, and two open-ended questions regarding (a) “What has been the most difficult or stressful event you have had to deal with?” and (b) “What has been the event that has most reinforced your pride in your professional behaviour?”.

## Analysis

The open-end questions were independently coded into themes developed by the authors; disagreement on which theme a response belonged to was resolved by counting partial agreement as agreement, given that overall, there was a high degree of concordance in coding (Gwet’s AC1 at least 0.8 for each theme); each open-ended response could belong to more than one theme. The authors classified themes in terms of their apparent relationship to factors in SDT that are hypothesized to either promote (themes of pride) or thwart (difficult or stressful events) intrinsic motivation.

We conducted exploratory principal components analysis on the measured mood disorders via HADS, history of anxiety and depression (and their treatment a year prior to the pandemic), having reported a distressing event or an event that reinforced pride in the profession. We determined the number of interpreted principal components using scree plots. We examined the association of “cases” of anxiety and depression in relation to themes from coded open-ended responses via multivariable logistic regression models, adjusting for demographics and health factors described above, as well as controlling for the profession and healthcare system. These yielded odds ratios (OR) and 95% confidence intervals (CI). Only effect estimates with p≤0.2 were interpreted. Heterogeneity of effects was evaluated using Wald-style test (Kaufman & MacLehose, 2013). All statistical calculations were performed in SAS v 9.4 (SAS Institute, Cary, NC).

## Results

### Descriptive statistics

We recruited 1,124 nurses and physicians for the study, most from TH (803 nurses, 174 physicians) rather than UMC (71 nurses, 74 physicians). More than a half, 621, shared the most distressing experiences with the rest not responding or indicating that none were applicable; physicians at UMC were the outliers with the lowest rate of report at 17 (22%). Almost a half, 510, reported some experiences related to reinforcing pride in their professions, with the physicians from UMC being an outlier at 11 reports (14%). Just over a third, 443, reported both stressful events and those that instilled pride in their professions.

Nurses were predominantly female (81%), White (89%), married or divorced (65%), aged 44 years on average (range 21-70), almost half had children under 18 years of age living at home (45%). Among nurses, 225 reported to have felt unwell since start of the pandemic and 16 reported a positive test for COVID-19; 45% had history of anxiety or depression with 20% reporting treatment for each a year before the onset of the pandemic.

There were more males among physicians (56%); they were more likely to be White (73%), married or divorced (68%), aged 50 years on average (range 24-75), almost half had children under 18 years of age living at home (49%). Among physicians, 43 reported to have felt unwell since start of the pandemic and 4 reported a positive test for COVID-19; 26% had history of anxiety or depression with 10% reporting treatment for each a year before the onset of the pandemic.

Exploratory principal components analysis suggests that there are four independent groups of nurses and physicians, accounting for 80% of common variance. There is a group with history of anxiety and depression that was also symptomatic during the pandemic who tended not to report either distressing or experiences they were proud of. The second group reported both distressing and proud events at the time of the survey but has neither history nor current symptoms of anxiety and depression. The third group was anxious and depressed at time of survey but had no history of these conditions and did not report any distressing or proudful experiences. The fourth group was characterized by both distressing experiences and being a case of depression, but low no anxiety scale and with no history of mood disorders. There results suggest that (a) persons with history of mood disorders are not the ones who are most likely to report events that either distressed or reinforced pride (groups 1 and 2) and (b) the history of mood disorders is not the sole driver of experiencing symptoms of anxiety and depression during the pandemic (groups 3 and 4). Eigenvectors and component factor profiles are given in **Supplemental Material A**.

### Logistic models of risk of anxiety and depression

Among nurses and physicians who reported distressful events, the rate of cases of anxiety was higher (33%, 202/607) than among those who did not (25%, 65/262) and the rate of cases of depression was higher (12%, 70/607) than among those who did not (9%, 23/262). Likewise, among those who reported incidences that reinforced pride in the profession the rate of anxiety was elevated (32%, 161/501) compared to those who did not (29%, 106/368) while the rate of depression was reduced (10%, 50/501) compared to those who did not (12%, 43/368). There was no evidence of multiplicative interaction between the reports of proudful and distressing experiences on the odds of either anxiety or depression. We only observed the evidence of distressing experiences elevating the odds of anxiety: OR 1.39 (95% CI: 0.96, 2.00) after accounting for healthcare system and profession.

We observed evidence of independent effects on depression, with *increase in odds* with report of distressing experiences (OR 1.48; 95%CI: 0.85, 2.55) and *decrease in odds* with reports of events that instilled pride in the profession (OR 0.71; 95%CI: 0.44, 1.14), after accounting for healthcare system and profession. The null hypothesis test for heterogeneity for the above two effect estimates yielded p=0.02.

The effect estimates for the above-mentioned logistic regressions models were not materially different after adjustment for other covariates (see Methods & materials).

Associations of specific themes of distressing experiences with anxiety and depression are illustrated in **Table 1**. The most commonly reported distressing experiences related to fear of infection (15%), concerns about use of and access to personal protective equipment (14%) lack of transparency in communications and administrative support (14%), and uncertainty (11%), each was reported by more than 100 respondents. When all factors were considered together, including adjusting for experiences that reinforced pride, the odds of anxiety was elevated among those who expressed worries about infecting immediate family (OR: 1.75; 95%CI: 0.89, 3.44), were dissatisfied with standards of patient care (OR: 3.09; 95%CI: 1.32, 7.24), and struggled with transparency of communications at work (OR: 1.50; 95%CI: 0.98, 2.29). Challenges with transparency of communication at work were also linked to elevated odds of depression (OR: 1.52; 95%CI: 0.85, 2.74). Furthermore, aggression from patients or their families (OR: 2.94; 95%CI: 0.66, 13.01) and feeling anger (OR: 2.66; 95%CI: 0.64, 11.07) where independently associated with depression after accounting for other factors.

**Table 1:**
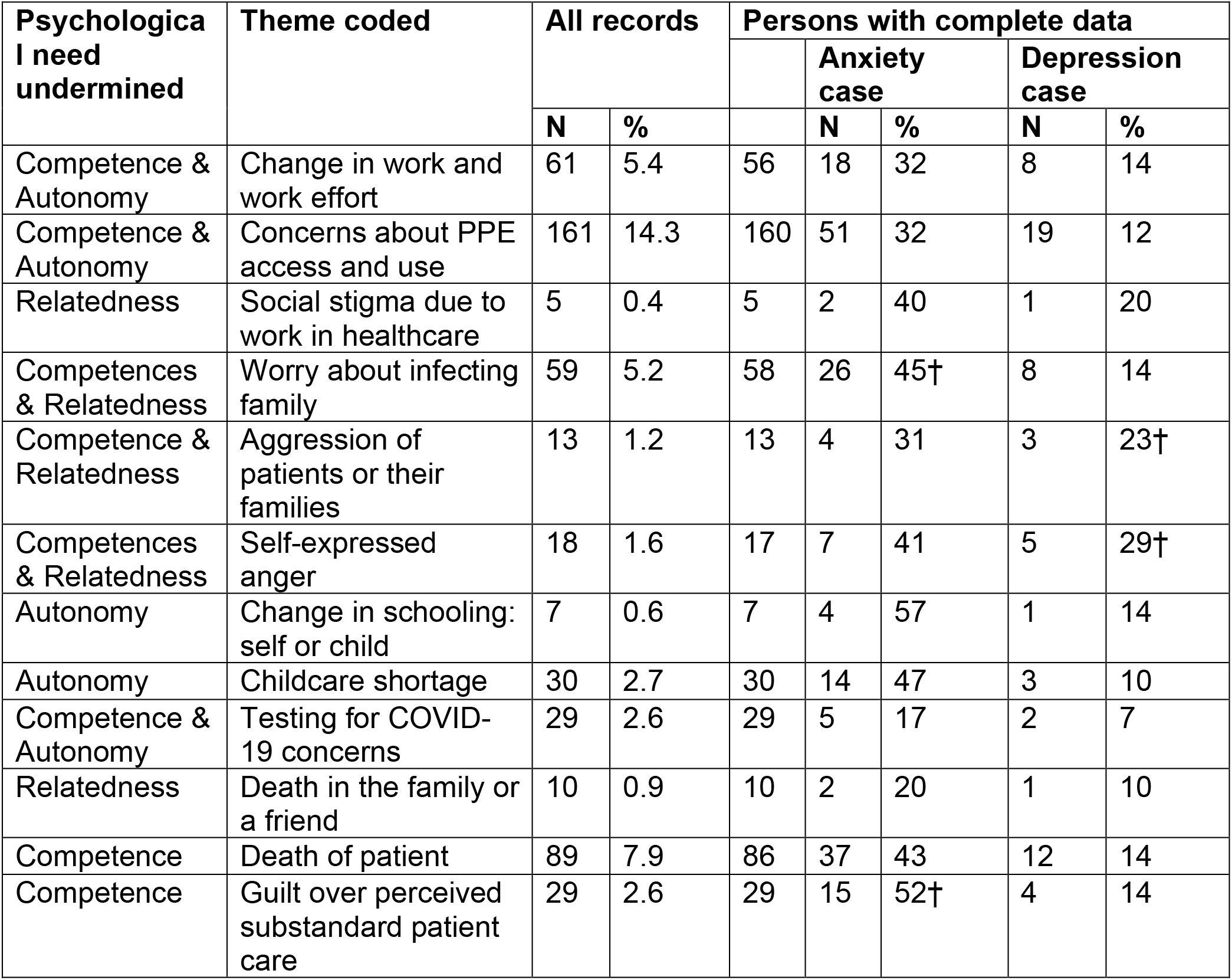
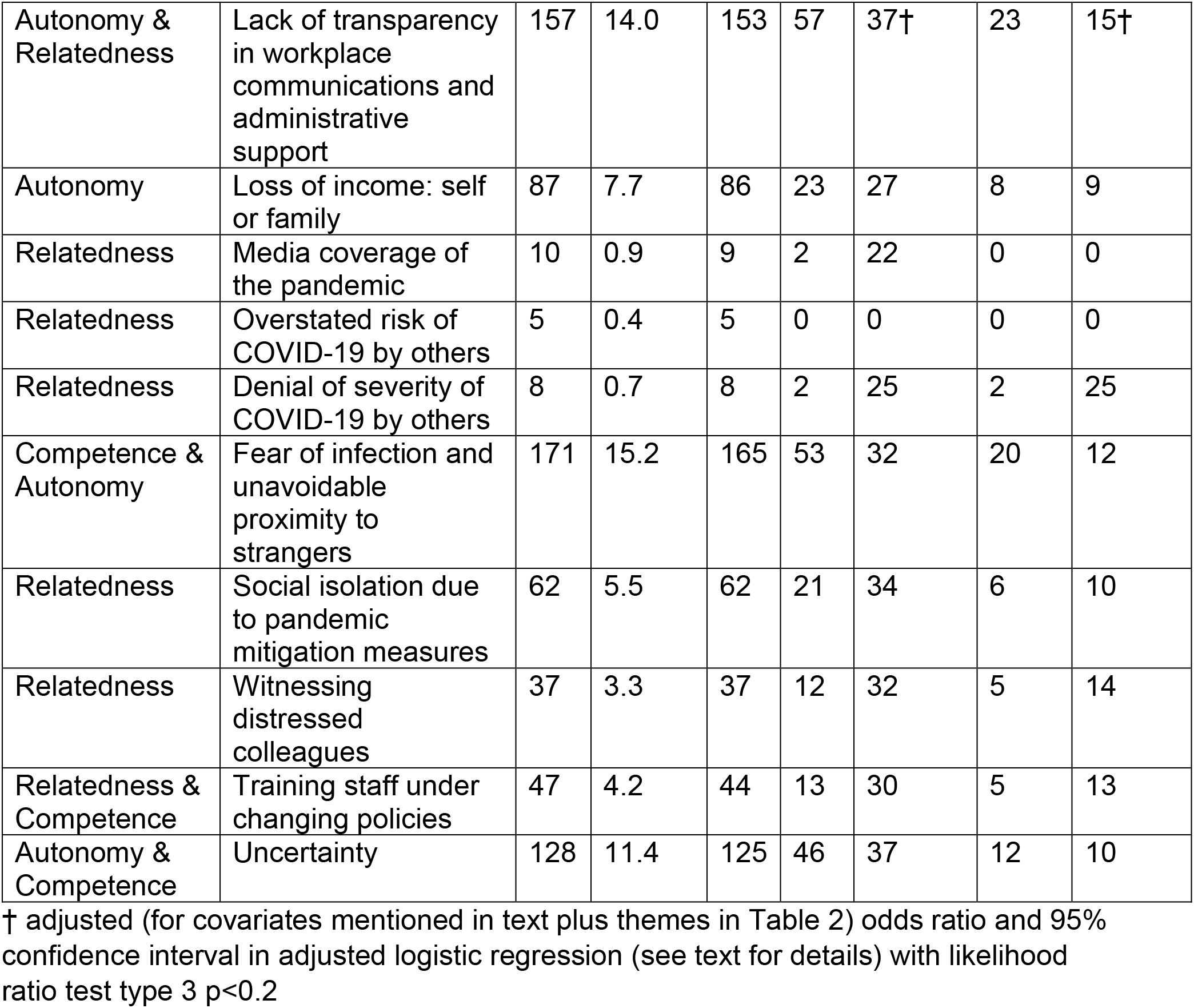
Distribution of responses of experiences related to most difficult experiences among 1124 physicians and nurses in relation the psychological needs from self-determination theory (STD) they undermined, and HADS scores >10 (cases of anxiety and depression) among 869 with complete data.

| Psychological need undermined | Theme coded | All records |  | Persons with complete data |  |  |  |  |
| --- | --- | --- | --- | --- | --- | --- | --- | --- |
|  |  | N | % |  | Anxiety case |  | Depression case |  |
|  |  |  |  |  | N | % | N | % |
| Competence & Autonomy | Change in work and work effort | 61 | 5.4 | 56 | 18 | 32 | 8 | 14 |
| Competence & Autonomy | Concerns about PPE access and use | 161 | 14.3 | 160 | 51 | 32 | 19 | 12 |
| Relatedness | Social stigma due to work in healthcare | 5 | 0.4 | 5 | 2 | 40 | 1 | 20 |
| Competences & Relatedness | Worry about infecting family | 59 | 5.2 | 58 | 26 | 45† | 8 | 14 |
| Competence & Relatedness | Aggression of patients or their families | 13 | 1.2 | 13 | 4 | 31 | 3 | 23† |
| Competences & Relatedness | Self-expressed anger | 18 | 1.6 | 17 | 7 | 41 | 5 | 29† |
| Autonomy | Change in schooling: self or child | 7 | 0.6 | 7 | 4 | 57 | 1 | 14 |
| Autonomy | Childcare shortage | 30 | 2.7 | 30 | 14 | 47 | 3 | 10 |
| Competence & Autonomy | Testing for COVID-19 concerns | 29 | 2.6 | 29 | 5 | 17 | 2 | 7 |
| Relatedness | Death in the family or a friend | 10 | 0.9 | 10 | 2 | 20 | 1 | 10 |
| Competence | Death of patient | 89 | 7.9 | 86 | 37 | 43 | 12 | 14 |
| Competence | Guilt over perceived substandard patient care | 29 | 2.6 | 29 | 15 | 52† | 4 | 14 |
| Autonomy & Relatedness | Lack of transparency in workplace communications and administrative support | 157 | 14.0 | 153 | 57 | 37† | 23 | 15† |
| Autonomy | Loss of income: self or family | 87 | 7.7 | 86 | 23 | 27 | 8 | 9 |
| Relatedness | Media coverage of the pandemic | 10 | 0.9 | 9 | 2 | 22 | 0 | 0 |
| Relatedness | Overstated risk of COVID-19 by others | 5 | 0.4 | 5 | 0 | 0 | 0 | 0 |
| Relatedness | Denial of severity of COVID-19 by others | 8 | 0.7 | 8 | 2 | 25 | 2 | 25 |
| Competence & Autonomy | Fear of infection and unavoidable proximity to strangers | 171 | 15.2 | 165 | 53 | 32 | 20 | 12 |
| Relatedness | Social isolation due to pandemic mitigation measures | 62 | 5.5 | 62 | 21 | 34 | 6 | 10 |
| Relatedness | Witnessing distressed colleagues | 37 | 3.3 | 37 | 12 | 32 | 5 | 14 |
| Relatedness & Competence | Training staff under changing policies | 47 | 4.2 | 44 | 13 | 30 | 5 | 13 |
| Autonomy & Competence | Uncertainty | 128 | 11.4 | 125 | 46 | 37 | 12 | 10 |

† adjusted (for covariates mentioned in text plus themes in Table 2) odds ratio and 95% confidence interval in adjusted logistic regression (see text for details) with likelihood ratio test type 3 $p < 0.2$

Associations of specific themes of experiences that instilled or reinforced pride with anxiety and depression are illustrated in **Table 2**. The most commonly reported proud experiences related to high quality of care provided (19%), teamwork (17%), and composure under stress (13%) each was reported by more than 100 respondents. When all factors were considered together, including adjusting for difficult experiences, the odds of anxiety was reduced among those who expressed pride about technological sophistication of care (OR: 0.38; 95%CI: 0.10, 1.41), altruism (OR: 0.65; 95%CI: 0.35, 1.21), and not becoming infected despite perceived exposure to the virus that caused COVID-19 (OR: 0.30; 95%CI: 0.09, 0.97). Support from community was independently associated with reduced odds of depression after accounting for other factors (OR: 0.47; 95%CI: 0.16, 1.34). Those who were proud of volunteering for hazardous tasks were at an elevated odds of both anxiety (OR: 1.70; 95%CI: 0.81, 3.55) and depression (OR: 2.47; 95%CI: 0.97, 6.29), after accounting for other covariates considered in our analysis.

**Table 2:** Distribution of responses of experiences related to pride in the profession among 1124 physicians and nurses in relation the psychological needs from self-determination theory they reflect and foster.

| Psychologic<br>al needs<br>fostered | Theme coded | All<br>records |  | Persons with complete data |  |  |  |  |
| --- | --- | --- | --- | --- | --- | --- | --- | --- |
|  |  | N | % |  | Anxiety<br>case |  | Depression<br>case |  |
|  |  |  |  |  | N | % | N | % |
| Autonomy | Adaptability and flexibility (self or colleagues) | 102 | 9.1 | 102 | 35 | 34 | 8 | 8 |
| Autonomy and Competence | Technological sophistication (inventiveness) in care for patients | 17 | 1.5 | 17 | 4 | 24† | 1 | 6 |
| Autonomy and Relatedness | Volunteered for hazardous tasks | 63 | 5.6 | 61 | 22 | 36† | 10 | 16† |
| Autonomy and Relatedness | Altruism | 74 | 6.6 | 74 | 21 | 28† | 10 | 14 |
| Competence | Composure under stress | 151 | 13.4 | 150 | 52 | 35 | 15 | 10 |
|  | Comforting dying patient | 19 | 1.7 | 19 | 10 | 53 | 3 | 16 |
|  | Not infected after exposure | 22 | 2.0 | 22 | 5 | 23† | 1 | 5 |
|  | Patients survive, quality of care | 216 | 19.2 | 212 | 66 | 31 | 21 | 10 |
| Relatedness | Community outreach by oneself | 19 | 1.7 | 19 | 8 | 42 | 3 | 16 |
|  | Support from community | 64 | 5.7 | 63 | 20 | 32 | 5 | 8† |
|  | Support from leadership | 43 | 3.8 | 43 | 12 | 28 | 2 | 5 |
|  | Teamwork | 196 | 17.4 | 189 | 61 | 32 | 18 | 10 |
|  | Thanks from patients | 42 | 3.7 | 42 | 17 | 40 | 6 | 14 |

† adjusted (for covariates mentioned in text plus themes in Table 1) odds ratio and 95% confidence interval in adjusted logistic regression (see text for details) with likelihood ratio test type 3 $p < 0.2$

### Themes of “What has been the most difficult or stressful event you have had to deal with?”

Frequencies and names of the coded themes among difficult or stressful events since the start of the pandemic are captured in **Table 1**. They also indicate our perceived association of these themes with autonomy-thwarting factors experienced by nurses and physicians. The nuances and meaning of the coded themes are illustrated below using the respondents’ own words, and are more fully presented in **Supplemental Material B**.

### Change in work or work-effort

Various changes in working conditions precipitated by the pandemic, not being planned events with buy-in from nurses and physicians, were stressful for most. The sheer increase in volume and pace of work was commonly mentioned: “Our days were non stop, not able to take an uninterrupted break.” On the other side of the spectrum, some changed involved “reducing the work force due to plummeting volumes”, explicitly related to furloughs and layoffs: “Furlough and lay-offs and the threat of no work -- VERY stressful.” For others, changes were more radical, involving alteration in the nature of work, e.g., towards primary clinical duties, and personal routines: “Being pulled to the ICU to care for Covid-19 patients without a full orientation and changing my schedule from day shift to evening and night. “ A related challenge that produced difficulties is the fluidity of procedures and demands: “the way things keep changing, it does not instill confidence.” For some, changes in work involved performing unfamiliar duties, while potentially endangering the usual (non-COVID-19) patients:

> “We have effectively taken on the role of dietary staff as well as housekeeping to limit people in each room (but without getting any additional staff). … we’re becoming very burned out. We’re told over and over that we need to re-use the same n-95s even though we know this is improper. They know its not safe, but we’re told to do it anyway. Its not fair. All of this is causing nurses to fall apart and this will cause a mass exodus from bedside nursing once all this is over.”

When such changes in the nature of work combined with poor communications and childcare obligations, difficult situations ensued: “Communication about daily changes and schedule was also lacking. I also have three children acclimating to on line school and was not always available to assist them with my schedule changes.”

Disruption of patient care not related to COVID-19 patients appeared challenging for care providers who were unaccustomed to not having answers for their patients: “It’s been logical that electives have been cancelled but difficult for those patients who are in pain and were looking forward to the surgery.” When change in work involved perceived reduction in quality of patient care, this proved stressful:

> “Not being able to offer touch, see facial expressions, or give a hugs has been a challenge.”

> “Video visits still very unwieldy and not easy to navigate for patients.”

Even if the type and patterns of work did not appear to materially change, the extra effort performing these duties during the pandemic-induced procedures proved a source of most notable stress for some, especially when dealing with critically ill patients and deaths, and some colleagues were not supportive: “We had multiple deaths each week and docs were more intense and demanding than usual causing me more stress to the point of crying.” Work effort also increased, causing distress, in cases where patients needed extra mental health support: “My patients’ anxiety and emotional response to COVID-19 and having to support them to ensure they do not become suicidal.”

### Economic insecurity, cancelled leaves

For some respondents, threats of loss of income to themselves or to colleagues proved demoralizing, a clear contradiction between messaging about the essential nature of work by nurses and physicians during the pandemic and the reality of economic conditions that restricted their practice:

> “Having work hours cut significantly has immensely elevated stressors. This has impacted my family financially, physically, and emotionally. In addition, not being able to provide nursing care in a pandemic has caused feelings of worthlessness.”

Some loss of income was seen as consequence of poor planning, causing needless increase in work effort:

> “Our department was ‘furloughed’ with the idea that once our census numbers were ‘back to normal’ so would our staffing. For weeks that was not the case. We were often working short with roughly the same number of patients.”

The issues related to change in policies affecting personal time off (PTO), one of the few things an employee typically controls, were upsetting to many, seen as means to save money at the expense of well-being of healthcare providers and patients, e.g.:

> “pto was taken away from staff members which was scheduled in November. PTO is not easily granted to begin with, now staff if[sic] having it taken away during a time when a mental health break is needed the most.”

### Unavoidable exposure to infected persons and fear of infection

Some nurses and physicians were distressed by their own “irrational fear of other” and “wondering if [they] will contract the virus no matter how careful”. Experiencing and anticipating risk of infection was reported among some of the most difficult experiences, with the underlying sense that infection control was not fully under one’s control, placing one’s family at risk: “I never-ever declined an assignment and lived in fear of infecting my family.” Concerns about testing for COVID-19 revolved around several themes, including “receiving patients … that were not tested and later turn out positive”, “patients lying during screening to reach face to face interaction with a provider”, “worrying about infecting others because I was not tested”, and “having … symptoms and waiting for my testing”. Some specific difficulties arose due to concerns about consequences of inaccurate test results, making a person “worried about the false negative tests”.

Given that “social distancing” was one of the proclaimed means to control the pandemic, when this was not enabled due to hospital policies or procedures, this resulted in frustration, likely fueled by a sense of powerlessness to avoid situations perceived as placing one at risk for infection: “All doors are locked and we’re supposed to be social distancing, but here we stand like a herd of cattle, at the few entrances to wait to get our temps taken.”

Becoming ill or exposed during the pandemic was recounted as one of the most difficult experiences of the pandemic, with the concerns centering on how this affected one’s family: “I contracted the disease and was very ill for 2 weeks, requiring a hospitalization. most stressed about how this affected my family”. When family members did contract COVID-19, a difficult situation ensured, especially when there appeared to be no additional support during the crisis: “My [spouse] was gravely ill with COVID and I had to care for both him and my children AND then return to work to care for my patients.”

### Personal protective equipment (PPE) and other infection control challenges

A matter closely related to risk of infection due to unavoidable contact with infected patients was the availability and utilization of PPE. Even possibly appropriate measures were a source of distress, like “constant” “… mask fit testing” and “… disinfecting”. Concerns about inadequate PPE were naturally intermingled with concern about infection, causing reports of distress:

> “Having to reuse the same N95 respirators every day, knowing that the more I use the same one, the less effective it becomes. I’ve also been working hands on with known Covid patients and the thought of putting the same dirty mask back onto my face every time disgusts me.”

Some reported difficulties that arose from trying to elicit collaboration of patients with the infection control measures: “patients not cooperating with prevention of spread.” For others, the concerns arose from the PPE and infection control measures adversely affecting communication that is integral to quality care: “The room has a huge loud fan for negative pressure you have a mask and a face shield on so not only can they [patients] not hear you at all but they can’t even read your lips.”

The effort involved in using PPE proved very onerous for many, including “wearing the masks for 12 hours”, “working 13 hours in N95 causing severe SOB, and dizziness”. A common related source of distress arose from having to re-use PPE, both due to discomfort, fear of shortages of PPE, and knowledge that this is leads to sub-optimal protection (i.e., against PPE training): “Having to reuse the same N95 respirators every day, knowing that the more I use the same one, the less effective it becomes.”

For some, the access to PPE was the major concern, combined with perceived lack of preparedness of the healthcare system to meet the challenges of a pandemic, and implied disregard for patient and staff safety: “The lack of adequate PPE putting my friends and coworkers at risk due to poor planning. To me this is inexcuseable[sic].”

A particularly difficult situations arose when it appeared that management refused to share the burden of risk with frontline workers who lacked PPE, actions perceived as cowardly and callous: “The [leadership team] let us go in and get contaminated but stayed in the hallway and watched.“

### Concerns about health of oneself and others, not related to contracting COVID-19

Being unable to receive proper medical care due to infection control protocols in place proved very difficult for some: “I had a scheduled appointment with my [specialist] and it was very upsetting and stressful that they wanted to reschedule it due to the sole fact that I am … working with COVID patients.” Some noted “lack of assistance getting access to mental health” as the causes of considerable distress, while others were affected by the increase in the mental health needs of their patients who exhibited “anxiety and emotional response to COVID-19” that led to “having to support them to ensure they do not become suicidal.” For a few, “unexpected death” of a family member during the pandemic overshadowed any other difficulties.

### Death of patients

For some nurses and physicians, the sheer rate of mortality was overwhelming: “5 deaths in one shift.” The circumstances of patients dying was among most difficult experiences, stressing the loneliness of these patients due to infection control protocols isolating them from their families repeated over and over again: “Seeing nurses so defeated in having to facetime with families to help the loved ones say goodbye…. This is heartbreaking”.

Some reported struggling with the sense of failure and futility of effort in trying to avert death of the COVID-19 patients, combined with belief that such patients could have received better care, including emotional support, even if they could not be saved: “Very difficult to watch a patient suffocate slowly over weeks only to die despite our best efforts. No family is allowed to visit. Horribly sad.” Even some veterans of the field who “can handle almost anything” found they “cried everyday” when dealing with dying COVID-19 patients who “tend to be hopeless” and were denied usual level of care by infection control protocols.

Some choices that nurses and physicians had to make, precipitated by infection control protocols, appear to be the stuff of ethical nightmares: “Patient likely going to die and having to limit which son will be able to remain.” A recurring, experience appeared to have been related to lapse in ethics of care due to lack of communication with families of patients: “… the DNR/AND is not honored. Families are not involved as much and do not understand what their loved one is being put through.”

### Perceived lapse (guilt over) in standards of patient care

Some nurses and physicians were burdened with “a lot of guilt” over belief that they did not provide the usual level of care: “I felt like I was killing people.” Some believed that they let their teams down: “Injuring myself … . I felt like I was letting my colleagues down by not being at work and going through all the stressful changes with them.” Those who had work hours reduced reported that “not being able to provide nursing care in a pandemic has caused feelings of worthlessness.” For some, concerns about quality of patient care appear to have been aggravated by breakdown in teamwork and fairness in allocation of responsibilities across a team: “Other people seem to be avoiding my ICU covid patients yet I am accused of not wanting to see them by a collegue. Meanwhile, my collegues I know aren’t seeing their patients and we are doing them a disservice … .”

### Social isolation

For some, the most difficult experiences of the pandemic arose from the loss of the usual social contacts. In many cases this was related to self-isolation for fear of spread of infection to family members at high risk from the virus, especially when combined with recognized mental health difficulties: “Dealing with my personal depression and anxiety, self isolation, not being able to see my mother in her SNF [skilled nursing facility]”. Isolation from family and wider community made coping with the pandemic more challenging for some, precisely because family, community and friends were the usual sources of support: “not being able to visit with my family and attend church.” For those who rely on co-workers for support and companionship, having to stay away from work for fear of spreading infection to vulnerable family members was among most difficult experience of the pandemic, identifying such circumstances as the cause of depression: “Staying home again after extended leave and being isolated from everyone has lead to depression - on top of the extreme fear of my … baby contracting the virus.” Working from home likewise resulted in erosion of social support from co-workers, such that “isolation due to working at home” was reported as being among the more difficult experiences whether the genesis was from guilt from not working to social isolation from friends.

In some instances, nurses and physicians felt ostracized by the community due to fear of contracting the virus, combined with perceived empty gestures of support and lack of tangible effort to make it easier for nurses and physicians to function: “I had to deal with friend, neighbors considering me a deadly weapon because I might be infected and a threat to them.” Difficult situations arose when nurses and physicians felt that their ability to communicate risk of the pandemic with their patients was superseded by misleading media coverage, resulting self-censorship, which is a form of social isolation: “It’s hard to do my job when families are constantly throwing cnn[sic] or google in my face! … This histeria[sic] has foced[sic] me to socially distant or i[sic] get social[sic] shamed.”

### Stressed colleagues

Give the importance of teamwork to successful provision of healthcare, perceived stress of co-workers proved to be among most difficult experiences for some: “Staff breaking down and crying; managing direct staff fears”. For some, breakdown in teamwork manifested in multiple ways, anchored in worries about risk of infection and unequitable workloads, and aggravated by lack of administrative support: ““Staff worried about PPE every shift. Upper management not supportive to staff. Physicians not going into patient’s rooms. Nurses taking on the roles of multiple jobs.”

Several nurses and physicians appeared find it difficult to deal with colleagues who appeared to not be bearing well under what was perceived as normal pressures of intensive care: “Mostly hearing co-workers ‘fishing for thank you’s’ [sic]. … being overly dramatic. … I wourk [sic] in the intensive care setting on a daily basis and this is not hard.” It appears that difficulties related to perceived “irrational fears” were conflated with frustration about how the pandemic was portrayed in the media and politicized: “The over the top societa[l], political response to a pedestrian disease”.

### Aggression and anger

While dealing with stressed colleagues was among most difficult experiences for some, encounters with agitated, angry and aggressive patients and their families was among the most adverse experiences of the pandemic for others, as it is a rare event to experience a patient’s or family member’s anger directed toward the nurse or physician:

> “Being yelled and screamed at by distraught, isolated suspected and confirmed Covid-19 patients because they did not feel I was moving fast enough … .”

> “Extremely agitated and combative behavioral health patients with Covid19 who cannot comply with instructions for safety of themselves and staff”.

Some nurses and physicians felt that their own anger was among the most difficult experiences of the pandemic, perhaps reflecting a negative emotion unusually directed toward patients or their families, presenting psychological needs for autonomy, competence and relatedness which were thwarted.

### Uncertainty

Uncertainty of the impact of the virus on one’s patients appears to have intermingled with sense that patients are not receiving the best possible care: “Every day feeling like I am failing my patients because we don’t know enough … .” The sense of uncertainty seemed widespread, akin to a sense of general apprehension: ““Feeling that I will encounter situations that I am not confident in my ability to manage.” The uncertainty about what holds true about the pandemic made it difficult for some to train staff, when official policies contradicted person’s understanding of the evidence, making it difficult to “coach … staff on policies/initiatives that [one does] not personally agree with.” Some nurses and physicians reported that “managing staff anxiety, fear” was particularly stressful in light of “knowledge deficits”. The constant changes, “at a daily and sometime hourly basis,” in the understanding of the pandemic at the leadership level precipitated stressful changes in procedures at the bedside: “One of most stressful situations I’ve had to deal with during the covid-19 pandemic was navigating the initial change in bedside care and policies.” For some, the diversity of views of the pandemic by the public and patients placed nurses and physicians in the crossfire of the competing narratives, such that the community’s uncertainty had an adverse spill-over effect on the frontline nurses and physicians: “interactions with people extremely paranoid about contracting the virus and interactions with people who don’t think the virus is real.” One respondent shared an experience of overcoming difficulties that arose from the stresses of chaotic initial period of the pandemic, by suggesting that they gained equanimity over time: “I feel like I grew out of that by continuing to work … “.

### Shortcoming in transparency at work

Some of the most difficult experiences related to “lack of communication”, “no answers, lack of answers, answers only to the individual asking”, including “not receiving information that the government and patients thinking [nurses and physicians] had access to testing both for the virus and antibodies before [nurses and physicians] did” and “no support from manager regarding how to find information or what to tell patients”. When practice guidelines appeared to make no sense and work-related requests were not perceived as having been dealt with rationally and respectfully, distressing situations arose: “… being told I did not need to wear PPE. After push back, getting the required PPE and being questioned why I needed them.” For some there was an overall sense of “the disorganization, poor communication, and frequent disrespect by [leadership/management]” which included perception of lack of concern for well-being of nurses and physicians: “leadership ignoring staff input … Being told we are ’heroes’ while being treated like second class citizens.” It was suggested that “an organized explanation, and a respectful conversation is always best even in a ‘crisis’ situation”.

### Lack of administrative support

An issue closely related to poor communication is perception of lack of administrative support, even among those who otherwise welcomed the challenges of the pandemic:

> “I enjoy a challenge, so the pandemic was an opportunity to prove that I can survive and be a positive support for others. The most stressful event so far is witnessing the poor leadership choices. Very disorganized. One minute they want us to do one thing, then the next it changes. Fragmented rules.”

When frontline works appeared to be overlooked in recognition by the management despite taking risks and speaking up in an attempt to remedy lapses in practice, this proved “stressful”: “we were constantly exposed and not recognized at all.” Worse still, “being disciplined for being frustrated with the increased work loaf [sic]” was named among most difficult experiences of the pandemic, leading some to sense that they are “unable” to perform their duties both safely and well. Some point to specific lack of support in reducing the uncertainty and improving patient care, namely “no support from manager regarding how to find information or what to tell patients.”

Perceived lack of empathy and “respectful conversation” with administration was led to “unnecessary frustration”: “the disorganization, poor communication, and frequent disrespect by my director and the command center.” When there was perception that rules were not fair, or not applicable to all, frustration arose due to: “… being ridiculed for wanting to follow set guidelines”. “Lack of empathy and understanding” from leadership appears to be recurring theme in situations that caused hardships to nurses and physicians, for example “when daycare closed and I had to figure out a schedule to still be able to work while my husband, who is also essential, was still able to work and our child was still cared for every day.” One of the expressions of this lack of empathy appears to have been seen in reports that “system is more worried about their bottom line than they are about patient safety”.

### Childcare

Loss of childcare and the resulting need to balance professional duties as an “essential” worker with family responsibilities was among most difficult experiences of the pandemic reported by those with children living at home: “Working full time … while the schools have been closed for the last two months, my [children] are at home …”. This was especially difficult when person was the only available caregiver: “My [spouse] was gravely ill with COVID and I had to care for both him and my children AND then return to work to care for my patients.”

### Themes of “What has been the event that has most reinforced your pride in your professional behaviour?”

Frequencies and names of the coded themes of the events since the start of the pandemic that reinforced pride in the profession are captured in **Table 2**. They also indicate our perceived association of these themes with autonomy-supportive factors experienced by nurses and physicians. The nuances and meaning of the coded themes are illustrated blow using the respondents’ own words, with detailed presentation of representative quotations in **Supplemental Material B**.

### Courage, composure under pressure

Given the anxieties and uncertainty of early days of the pandemic, it is not surprising that individuals who felt that they conquered their fears and remained on the job were proud of the fact: “… I haven’t allowed COVID to overtake my life with fear/anxiety”. Some were proud not only of the fact they continued to work but that they could say: “I still love the work I do”. The pride in overcoming personal fears was related to putting the needs of patients above personal risks: “Showing up at work everyday [sic], odd shifts despite the hardships of felling unprepared without all of the answers and constraints on family life (daycare closed, worried about my family, not seeing friends and neighbors).” For some, the pride in steadfastness of nurses and physicians intertwined with pride in effective teamwork: “The resolve of the … doctors and nursing staff … . There is a collegial feeling of purpose that is palpable but hard to articulate.” Some experiences that instilled pride combined the sense of being up to the challenge of the pandemic as the team, while receiving little support from administrators: “We succeeded in spite of administration. Not because of it.” This pride in autonomous professional success is summarized succinctly as pride due to “relying On myself”. Some noted that they were proud not in just how well they worked but by their professional growth: “My confidence has grown throughout this whole experience … .”

### Volunteering for hazardous tasks and altruism

One clear example of courage under duress was expressing in volunteering for tasks perceived to be hazardous, an understandable source of personal pride: “I will see covid patients to keep another provider, at higher risk that I, from seeing them”. Some nurses and physicians were proud of doing what they thought was right for their patients and colleagues, despite personal risks: “Being able to be there for the community and treat patients while risking my own self and family’s health.”

This did not necessarily relate to volunteering for hazardous duties but taking extra effort to do what one thought was right, even when there was perceived lack of support from the administration for such acts: “Ultimately I had to use community resources and also my own money to buy [N95 masks and face shields] from to community also, but i’m[sic] proud I kept my staff safe. This was both very stressful and proud moment.” Altruism and courage displayed by others was a clear source of pride about the profession overall: ““Everyone did what they had to do during the early aids epidemic before we knew what was going on some docs refused to treat patients we had none of that this time.”

### Quality of care, positive outcomes

Among most common experiences that reinforced pride in the discipline among nurses and physicians was the quality of care they provided, exemplified by positive outcomes: “the few who have been the most sick with covid and survived”. Beyond survival, specific aspects of patient care that were highlighted among those that instilled pride included “successfully educating patients about what we currently know and easing their fear and anxiety”. Pride in compassionate “high quality of empathetic, therapeutic care” was abundantly evident, especially needed due to isolation of patients precipitated by infection control measures: “being able to make these isolated pts smile …”. Success in infection control was a source of pride: “Knowing that I have come to work through this all and have not contracted Covid.” Some shared that they were proud of being able to help others provide the high-quality care: “That my former students at the bedside are setting examples of high quality, compassionate, and safe patient care”.

Examples of assertiveness and successful advocacy on behalf of the patients were a source of pride, resonating with themes of pride in autonomous action: “I am proud of my ability to advocate for suffering patients to get physician orders for oxygen therapies and morphine drips.” A related source of pride is how nurses and physicians adapted to caring for patients, growing in both confidence and skill required to practice their profession under the pandemic’s constraints; there was a sense that this is what the profession was meant to do: “We were the first COVID unit. Watching the nurses go from being uncertain, scared to totally embracing it and doing an excellent job caring for these patients. Putting patients first. This is our calling, it’s why we are nurses.” Some shared that their pride rested on having provided quality care despite lack of recognition and support: “It hurts that we were never recognized for the hard work we did, but we didn’t do it for the recognition anyway, but for our patients.” For many of the respondents, pride arose simply from doing their work despite pandemic: “I am still providing the same quality of care that I did before the pandemic and that is what I am proud of.”

### Comforting dying patients

Despite death of patients being named among most difficult experiences by nurses and physicians, the manner they handled death of patients was a source of pride for many: “None of My patients died alone. I was there for each of them.”

### Innovation and technological sophistication

Some were clearly proud of their own inventiveness in overcoming challenges posed by the infection control protocols, reporting that they were proud of: “coming up with out of the box ideas … to facilitate better communication with patient and caregiver.” The fact that developing new way of providing care necessitated initiative due to (perceived) lack of organizational support was a source of pride for some: “Working without any instruction on COVID patient care from hospital administration”.

### Community outreach beyond clinical duties and support from community

Some respondents were proud of the fact that they contributed to management of the pandemic outside of their immediate clinical duties: “trying to be the middle ground, voice of reason and making sure I have researched facts I provide to friends/family/patients.”

For some, pride stemmed from recognition of nurses and physicians by the wider community, which included both expressions of gratitude but material support: “The outpouring of community support, donations, thank yous[sic], and the complete strangers that were kind enough to make scrub hats for us.” Recognition of difficulties faced by nurses and physicians by media appeared to be an important part of instilling a sense of pride: “I appreciate the truth about the lack of crucial medical supplies and PPE … the news reporting how medical staff and nurses are treated when we try to ourselves safe and healthy by advocating for ourselves and speaking up for what is happening that is not right.” Expressions of gratitude and concern that had a personal touch appeared to be appreciated such as when a formed patient called: “… to ask if the nurses are ‘OK’ because he was worried … .” Expressions of support and gratitude from family members who thereby assumed person risk of infection were likewise a source of pride: “My family supporting me in continuing my job as a nurse, despite being away from home, while friends were telling me to get out of nursing due to COVID.”

### Adaptability and flexibility

Nurses and physicians reported to be proud of how they and their colleagues adapted to the challenges of the pandemic: “Everyone’s resourcefulness and support in coming up with ways to continue to take care of pts in ways that keep exposure to a minimum.” For some, adaptability that they were proud of appeared to be facilitated by effective teamwork: “Teamwork, sacrifice, and agility of each member of our staff to re-organize and implement provision of care while keeping patients and staff safe as possible.”

### Revealing leadership qualities

Some we proud of how they led their teams: “I took it upon myself to guide and take care of my team during time where office manager lacked decision making and / or communication with staff”. Some found effective leadership displayed by others a source of pride for the profession as a whole: “… the course shown by some nursing, physician and several other staffs.” Those in the leadership positions appreciated being recognized when they proved effective: “Having the employees at the end of our conversations thank me for the help …”.

### Teamwork

Teamwork was a source of pride in how some nurses and physicians comported themselves, despite fears and uncertainties. This is exemplified by a report “that nurses and other health care providers, even when scared without having information, still came to work and did their job for their patients.” The sense of *esprit de corps* and focus on the mission to serve patients despite personal risk is clearly articulated in this narrative:

> “The professional behavior that has most reinforced my pride in healthcare has to recognize the human spirit within all health care workers. For me, watching nurses and doctors work expertly toward one mission, saving the life before you while keeping all stakeholders involved in the care safe from harm and illness. I admire their ability to set aside their fears and go into action, regardless of their internal insecurities.”

The pride in teamwork also arose because it entailed peer support: “Happy with support of co-workers at coming together to work in a pandemic.”

### Gratitude from patients

The sense of professional pride was reinforced when patients expressed gratitude: “compliments from my patients, when they tell me they appreciate me listening to them and taking the time needed to take great care of them”. It appears that gratitude from some patients and their families was related to both quality of care and recognition of personal risks that nurses and physicians endured in order to care for the patients:

> “A woman with [condition that is not COVID-19 redacted] said to me with tears in her eyes after her surgery, ‘Thank you for being here for me. With all that is going on, I was so worried that there wouldn’t be anyone to take care of me.’”

### Recognition from administrators and leadership of healthcare systems

Nurses and physicians who believed to have been appreciated by their employers and leaders within their organizations tended to report this as one of the experiences that reinforced pride in their professions: “Receiving a quality award for exceptional performance”. When organizations did not come across as divided between nurses and physicians and administration, there was a reason to report this as a proud achievement: “The love, support and spirit of my nurses, first responders, and hospital administration.” For some nurses and physicians, the pride in their professions was reinforced by acts of advocacy by those in a position to do so: “my union’s persistence”.

### Not proud

Some respondents elected to tells us why there were not proud of their profession, even though we did not inquire about this. Some of this relates to regret of not having done enough: “I should have tried harder to get the necessary resources … I should not have let the counseling deter me for advocating for myself and my colleagues.” The sense of not being able to advocate for the best handling of the pandemic appears to be a common them among those who appear to feel less than proud about their profession’s and their personal contribution during pandemic, with the source of frustration aimed at government and hospital leadership: “Government is causing the numbers to be inflated because they are paying hospitals for every patient diagnosised[sic] with CoVID[sic], even more if they are intubated or die as the diagnosis.” Others expressed dismay at how healthcare systems reacted, conveying a sense of futility and apathy in light of a contradiction between known best practices and demands of administrators: “I try to keep it together for my patients, but who cares about PPE regulations …We know this is wrong, but our administration tells us to do it anyway- so why try?” Some responses appear to capture a sense of exhaustion, burn-out: “I don’t[sic] have any feelings any more to my profession. … Having feelings is not much use.”

It appears that for some respondents pride in the profession depended on action of external forces and how they were treated rather than intrinsic values or achievements: “Pride is out the window. We have been treated increasingly worse as this rolls on. Most staff are barely hanging on and several have quit.” It would appear that for some the value of recognition within organizations and faith in rational actions in the best interest of patients outweighs positive impact of support from community at large: “Media calls us heroes while leadership treats us like an expendable commodity. … MANY patients and staff exposed due to incompetent decisions … .”

## Discussion

We believe that our contention that autonomy-supportive experiences lead to positive perception of the pandemic and better mental health is borne out by thematic analysis and association of the elicited themes with measures of anxiety and depression. For example, we observed that experiences that undermined the sense of competence, such as in performance of clinical duties, were associated with anxiety. Breakdown in communications and administrative support at work appeared to undermine the sense of autonomy (e.g., instructions on what to do constantly changing and apparently without justification) and relatedness (e.g., a sense of administration working against, not with nurses and physicians), and was correlated with both greater anxiety and depression. Pride in technological sophistication and inventiveness are clear indications of sense of autonomy and competence which lessened anxiety. A sense of pride that emanated from community support can be seen as support for sense of relatedness and was protective against depression. Against our hypothesis of theoretical associations, volunteering for hazardous tasks, which can be seen as expression of both autonomy (i.e., person volunteered) and relatedness (e.g., such acts were motivated by desire to protect co-workers more at risk from COVD-19) was associated with heightened anxiety and depression. It is perhaps persons who are proud of such experiences who need to be targeted for mental health supports, in addition to those who struggled with distressing events. Overall, any distressing events were related to both anxiety and depression but reports of events that instilled pride counter-acted risk of depression (i.e.. feeling good about one’s individual clinical acumen can be beneficial). This again suggest that *fostering a sense of pride in how one comports themselves may an effective means to safeguarding against depression and anxiety during disasters* such as COVID-19 pandemic.

Our findings must be placed in the context of the US healthcare system. Nurses and physicians in the US are trained to be independent practitioners. This means they assess patients, make decisions as to what care or treatment is called for, and then develop a plan with the patient and their family. When the COVID-19 pandemic arrived in the US, as these nurses and physicians struggled to develop a plan for care, the promising practices of treatment were in development and in flux from one day to the next according to the CDC recommendations and ever-changing policies of the hospital administrations. This caused undue stress on the providers of healthcare, who were used to *independently developing and implementing plans of treatment* without interference from local, regional, or national hospital administration. With the national management of the COVID-19 pandemic, this was not the case, and we hypothesize that this led to the frustration, anger, disappointment, anxiety, and lack of individual pride in practice, which we captured in the data. For the future, policymakers may respect practitioners, encourage them to innovate during crises, allowe them to feel proud about their work as a coping mechanism, and not inconsequentially, may develop a clinically relevant practice, such as posturing patients, became during the pandemic.

The narratives captured patient experiences during COVID-19 pandemic, which was not among our aims, and concerns of nurses and physicians about quality of care (correlated with anxiety). *Did nurses and physicians advocate for their patients when asked about their own experiences of the pandemic?* It can be inferred from distressing experiences of nurses and physicians, that infection controls that prevented families from being with dying patients should have been altered to be more humane. Likewise, the narratives imply a question as to whether mental health support and communication protocols were in place to supply best possible patient care. We think that these results can shape policies that are beneficial to both nurses and physicians and patients.

Themes of most difficult events among nurses and physicians overlapped with those elicited in a sample of general population in Philadelphia, where TH is located, over a similar time period as this report (Burstyn & Huynh, 2021). Specifically, the themes shared across samples were economic woes, disruption of working lives, childcare struggles, worries about health (including contracting COVID-19), uncertainty, media coverage, and frustration with government response to the pandemic. Themes associated with mood disorders that appear common to the two samples were worries about contracting COVID-19 and experiencing inconsistent messages and poor support from those perceived to be positions of power (respectively: hospital administration vs. government), implying that they are not specific to nurses or physicians, and may be related to universal psychological needs articulated by the SDT.

Our work suffers from numerous limitations arising from cross-sectional design, even though we did control for health history and demographics. Notably, principal component analysis shows that both difficult and pride-full experiences are not exclusively clustered among persons with history of mood disorders. However, all data is self-reported, thus being subject to biases from social desirability and correlated errors. Lack of responses in shared narratives cannot be interpreted as absence of relevant events and our conclusions are thus tempted by bias due to willingness to share experiences. Some of the experiences, such as those arising from management environment and economic situation must have been shared by all respondents within healthcare systems and professions, but only reported by some. Therefore, our conclusions are limited to perception of events and willingness to share them; we mitigated bias from shared working conditions by controlling for healthcare system and profession in statistical models. We lacked some of the information that would have been helpful in interpreting data, such as availability and utilization of mental health supports. We only studied two healthcare systems and struggled with (typically) low participation rates, undermining generalizability of the findings.

Despite limitations, we believe that our work offers some valuable insights and can help manage mental health challenges experienced by nurses and physicians during response to epidemics. For example, the elicited narrative themes of the most difficult or distressing events and moments that instilled pride in the professions can be the foundation of a survey instrument on perception of pandemics. This may assist in monitoring wellbeing of nurses and physicians during response to emergencies and obtaining their buy-in with changes in care provision. It is plausible that feedback from nurses and physicians to leadership that is inherent in autonomy-supportive buy-in would improve patient care, given that nurses and physicians were proud to share their innovative solutions to challenges of patient care during the pandemic. We note that some themes are not specific to infectious disease outbreaks but rather speak to autonomy-supportive workplace practices and leadership in general. Such practices and leadership can be fostered within healthcare systems and evaluated. In particular, autonomy-supportive leadership is suggested to be related to improved well-being in a recent meta-analysis (Slemp et al., 2018). It was discussed how leaders among nurses can improve mental health of other nurses during COVID-19 pandemic (Hofmeyer & Taylor, 2021). Additionally, it is indicated that during COVID-19 pandemic specifically, autonomy-supportive environment anchored in perceived social responsibility of the employer (but not extrinsic motivation) produced higher performance among employees, presumably in part by protecting their mental health under duress of pandemic-precipitated disruptions (Camilleri, 2021).

## Conclusion

Our findings document the broad spectrum of difficult and positive experiences of some nurses and physicians during the first wave of COVID-19 pandemic. We offer insights into how to monitor healthcare workers’ well-being during pandemic and provide evidence that autonomy-supportive policies may foster well-being under duress.

## Data Availability

Access to data is restricted by IRB but the essential elements of qualitative data are contained in the manuscript.

## Supplemental Material A

**Table A1:**
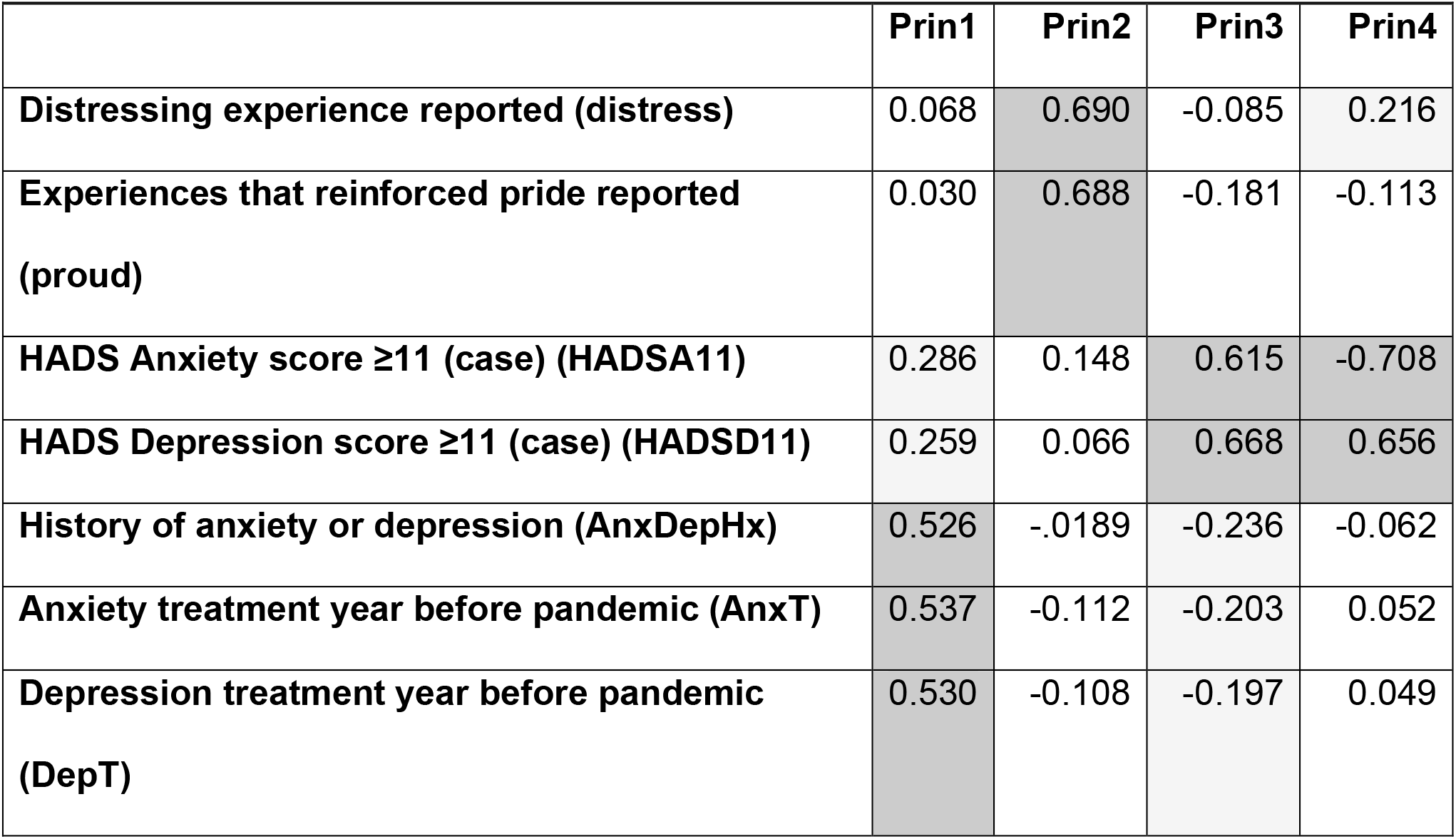
Eigenvectors of principal components (Prin) examining the inter-relationship of binary reports of distressing experiences, experiences that reinforced pride in the profession, Hospital Anxiety and Depression Scores (HADS) at the time of data collection that indicate case of anxiety or depression, history of anxiety or depression prior the COVID-19 pandemic, and reports of treatment year before pandemic for anxiety or depression.

**Figure A1:**
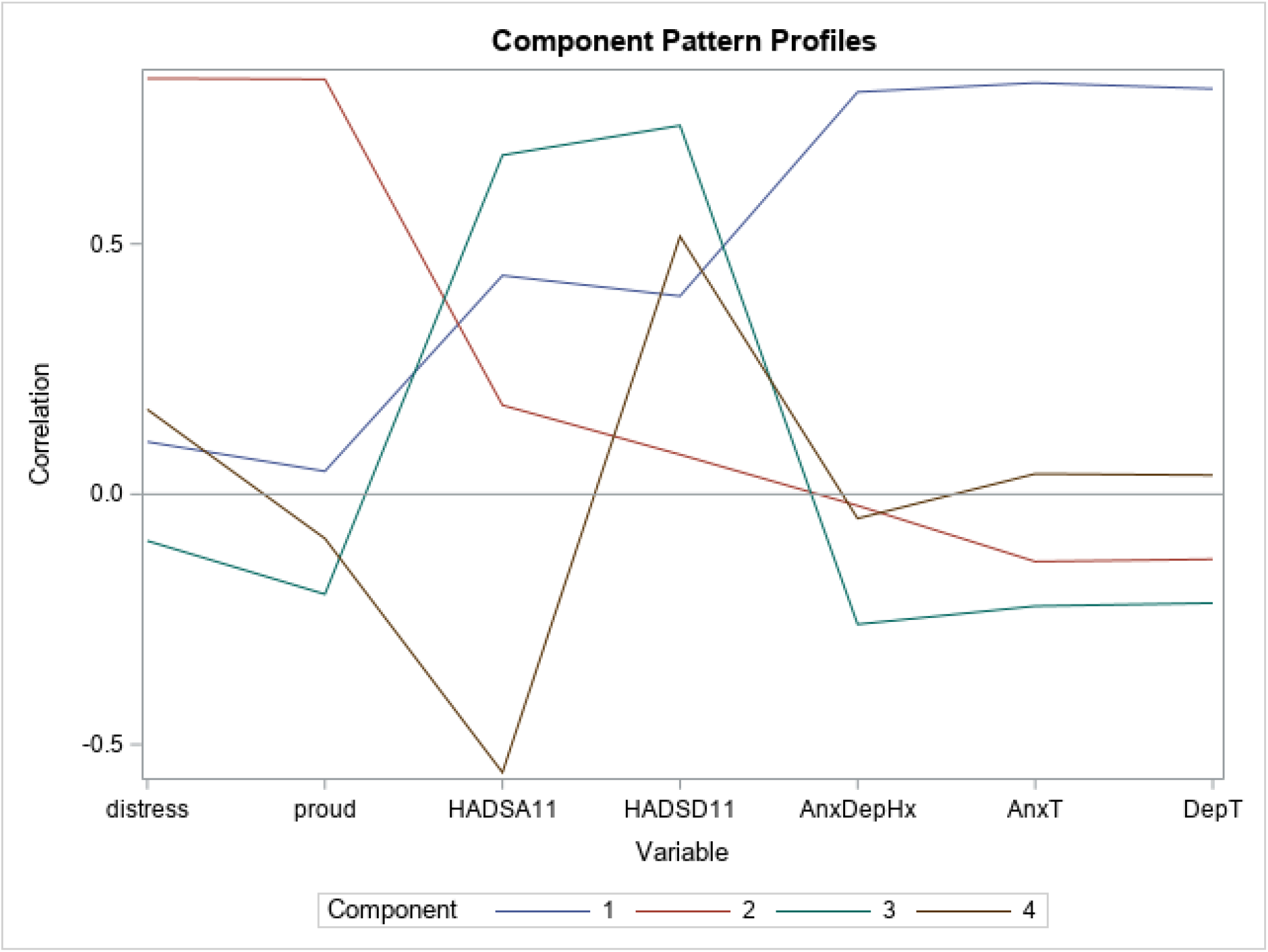
Component factor pattern profiles from the principal components analysis with eigenvectors from Table A1 (se for variable definitions).

## Supplemental Material B

**Pride and difficulties among nurses and physicians during the first wave of COVID-19 pandemic in two US healthcare systems: Mixed methods analysis of a cross-sectional survey: Detailed presentation of thematic analysis**

### What has been the most difficult or stressful event you have had to deal with?

Frequencies and names of the coded themes among difficult or stressful events since the start of the pandemic are captured in **Table 1**. They also indicate our perceived association of these themes with autonomy-thwarting factors experienced by healthcare workers. The nuances and meaning of the coded themes are illustrated blow using the respondents’ own words.

### Workplace factors: Change in work or work-effort

Various changes in working conditions precipitated by the pandemic, not being planned events with buy-in from healthcare workers, were stressful for most. The specific changes that were mentioned varied. The sheer increase in volume and pace of work was commonly mentioned:

> “Our days were non stop, not able to take an uninterrupted break.”

> “taking care of sicker patients without an increase in staff support”

> “We flexed our shifts … to limit our exposure. Our office WAY too small for that, the weeks you were working, you were HAMMERED.”

On the other side of the spectrum, some changed involved “reducing the work force due to plummeting volumes”, explicitly related to furloughs and layoffs: “Furlough and lay- offs and the threat of no work -- VERY stressful.” For others, changes were more radical, involving alteration in the nature of work, e.g., towards primary clinical duties, and personal routines:

> “Change in my work responsibilities from education to bedside nurse. … Have not worked nightshift in over 20 years.”

> “Going to medical unit with all covid patients to perform care I hVE NOT GIVEN IN 35 YEARS.”

> “Being pulled to the ICU to care for Covid-19 patients without a full orientation and changing my schedule from day shift to evening and night. “

A related challenge that produced difficulties is the fluidity of procedures and demands: “the way things keep changing, it does not instill confidence.”

For some, changes in work involved performing unfamiliar duties, while potentially endangering the usual (non-COVID-19) patients:

> “Not being allowed to do my job because of low census in outpatient area and being told I have to work in inpatient to help out and potentially be exposed to COVID-19 patients and then potentially bringing it back to my outpatient [redacted] patients who are at high risk”

When such changes in the nature of work combined with poor communications and childcare obligations, difficult situations ensued:

> “Communication about daily changes and schedule was also lacking.I also have three children acclimating to on line school and was not always available to assist them with my schedule changes.”

> “Poor communication throughout the hospital organization regarding safety and PPE use. One shift surgical masks were acceptable the next shift needed N95. Rules and safety measures were constantly changing and there was minimal communication.”

Disruption of patient care not related to COVID-19 patients appeared challenging for care providers who were unaccustomed to not having answers for their patients:

> “It’s been logical that electives have been cancelled but difficult for those patients who are in pain and were looking forward to the surgery. Many have reached out to me for support and answers. I don’t have answers- no one does.”

> “watching patients not be cared for in routine ways that would have been standard pre COVID has been particularly troublesome.”

> “Making difficult decisions regarding treatment with no or limited evidence.”

When change in work involved perceived reduction in quality of patient care, this proved stressful:

> “… we are very hands-on with our patients. Not being able to offer touch, see facial expressions, or give a hugs has been a challenge.”

> “Patient’s are typically elderly so their often hearing impaired. The room has a huge loud fan for negative pressure you have a mask and a face shield on so not only can they not hear you at all but they can’t even read your lips.”

> “Since I work in the NICU mainly, mothers and fathers cannot visit their babies as often as they would before.”

> “Video visits still very unwieldy and not easy to navigate for patients.”

Even if the type and patterns of work did not appear to materially change, the extra effort performing these duties during the pandemic-induced procedures proved a source of most notable stress for some, especially when dealing with critically ill patients and deaths:

> “… the nursing unit was converted to all negative pressure rooms with the use of HVAC air filters in the room. They were very loud and big. The sound of the air filter made it VERY difficult to hear each other, there was not efficient close loop communication. It was a very hectic stressful rapid response since we were unable to hear each other.”

> “We had multiple deaths each week and docs were more intense and demanding than usual causing me more stress to the point of crying.”

Work effort also increased, causing distress, in cases where patients needed extra mental health support:

> “My patients’ anxiety and emotional response to COVID-19 and having to support them to ensure they do not become suicidal.”

### Workplace factors: Economic insecurity, cancelled leaves

For some respondents, threats of loss of income to themselves or to colleagues proved demoralizing, a clear contradiction between messaging about the essential nature of work by healthcare workers during the pandemic and the reality of economic conditions that restricted their practice:

> “The potential of losing my job and being able to provide for my family. As a nurse this is not a fear I really ever thought I would have to worry about.”

> “Decrease in work/paid hours and working in areas of hospital I am unfamiliar with”

> “I will not be able to pay the myself and office staff that have been on the front lines with me”

> “I was fine until yesterday, when [healthcare system name redacted] laid off [redacted]000 employees. I have extreme distress that once again, the little guy pays the financial price for things beyond our control. I feel guilty that I still have a job and I am worried that now I don’t have enough support staff to do my job effectively.”

> “The Hospital system is more worried about their bottom line than they are about patient safety and it shows: they’re cancelling all flex time and PTO for anyone not working on a COVID unit so that they don’t have to pay a flex time shift differential, but all staff who are full time on a COVID unit have all of their PTO cancelled, and we’re becoming very burned out.”

### Workplace factors: Unavoidable exposure to infected persons and fear of infection

Some healthcare workers were distressed by their own “irrational fear of other” and “wondering if [they] will contract the virus no matter how careful”. Experiencing and anticipating risk of infection was reported among some of the most difficult experiences, with the underlying sense that infection control was not fully under one’s control, placing one’s family at risk:

> “working with COVID positive patients for 12 hour shifts and walking out of the hospital wondering if I’ve contracted the virus and who I’m giving it to. I felt like a ticking time bomb”

> “I never-ever declined an assignment and lived in fear of infecting my family.”

Concerns about effectiveness of testing for COVID-19 were at the root of some healthcare workers’ fear of infection, and in some cases intertwined with having to also deal with angry and frustrated of patients:

> “Hospital was behind in having COVID-19 testing site available and turning away multiple patients because the[y] did not have a fever of 100.4 F or higher to later find out, not everyone experiences a fever but can still be positive for the virus. This put people at risk of spreading the virus not knowing if they had it or not, since they were refused testing. Also, patient’s … yelling at nurses/staff and demanding to be tested for COVID-19 virus while nurses/staff continue to work & put ourselves at risk for contracting the virus.”

Concerns about testing for COVID-19 revolved around several themes, including “receiving patients … that were not tested and later turn out positive”, “patients lying during screening to reach face to face interaction with a provider”, “worrying about infecting others because I was not tested”, and “having … symptoms and waiting for my testing”. Some specific difficulties arose due to concerns about consequences of inaccurate test results, making a person “worried about the false negative tests and that people in the surrounding areas are not using proper social distancing and wearing masks”.

Distress at the risk of being infected due to inadequate testing was combined for some with self-isolation from support network and inadequate communication regarding infection risk:

> “Every day feeling like I am failing my patients because we don’t know enough, don’t have adequate plans in place, are not allowed to test in our outpatient office so we can’t give people good information or the care they desire. That I will infect my family, that I can’t see my parents or any of the rest of my family and friends because I am exposed.”

> “Being separated from my family to keep them safe. Stayed in hotel after shifts at hospital. When last shift completed I stayed additional 10 days before coming home.”

Given that “social distancing” was one of the proclaimed means to control the pandemic, when this was not enabled due to hospital policies or procedures, this resulted in frustration, likely fueled by a sense of powerlessness to avoid situation perceived as placing one at risk of infection:

> “Getting into the hospital. All doors are locked and we’re supposed to be social distancing, but here we stand like a herd of cattle, at the few entrances to wait to get our temps taken. There has to be a better way!”

Evidence of exposure to the virus from unknown sources was among most difficult reported experiences, such as “testing positive for antibodies with no patient contact”.

Becoming ill or exposed during the pandemic was recounted as one of the most difficult experiences of the pandemic, with the concerns centering on how this affected one’s family:

> “I contracted the disease and was very ill for 2 weeks, requiring a hospitalization. most stressed about how this affected my family”

When family members did contract COVID-19, a difficult situation ensured, especially when there appeared to be no additional support during the crisis:

> “My [spouse] was gravely ill with COVID and I had to care for both him and my children AND then return to work to care for my patients.”

### Workplace factors: Personal protective equipment (PPE) and other infection control challenges

A matter closely related to risk of infection due to unavoidable contact with infected patients was the manner of utilization of PPE. Even possibly appropriate measures were a source of distress, like “constant” “… mask fit testing” and “… disinfecting”.

Concerns about inadequate personal protective equipment (PPE) were naturally intermingled with concern about infection, causing reports of distress:

> “I must come to terms with the fact that I have been caring for this patient in close contact for hours or days without full PPE prior to them showing symptoms. I worry that I will get sick, and that I may spread it to my coworkers and family.”

Some reported difficulties that arose from trying to elicit collaboration of patients with the infection control measures:

> “patients not cooperating with prevention of spread (they will not use masks, limit their visitors, not telling us they are positive or have symptoms).”

For others, the concerns arose from the PPE and infection control measures adversely affecting communication that is integral to quality care:

> “Patient’s are typically elderly so their often hearing impaired. The room has a huge loud fan for negative pressure you have a mask and a face shield on so not only can they not hear you at all but they can’t even read your lips.”

The effort involved in using PPE proved very onerous for many, including “wearing the masks for 12 hours”, “working 13 hours in N95 causing severe SOB, and dizziness”, leading some to suggest that PPE was causing more harm than good:

> “Wearing an N95 for 12 hours makes you irritable. It gives you chest pain, nausea, and a headache. Not to mention the marks on your face. You wear the same disgusting one day in and day out. You develop acne.You are absolutely miserable driving to work and almost hoping you get sick so you are able to take some time off.”

> “Having to wear a mask all the time as I am claustrophobic.”

> “I am worried about wearing the same mask for 36 hrs or 3 shifts. I’m worried about the sterilization Of masks and what those chemicals will do to me.”

A common related source of distress arose from having to re-use PPE, both due to discomfort, fear of shortages of PPE, and knowledge that this is leads to sub-optimal protection (i.e., against PPE training):

> “Wearing an N95 for 4 weeks, and then another 3 weeks at a time is dangerous to staff health breathing in CO2 for extended hours and far beyond general recommendations for mask wearing. the fear of discarding the n95 mask and not being able to obtain a new one was a challenge.”

> “The need to wear surgical and N95 masks repeatedly when it was so engrained in you to change them between every surgical case and to remove them from your body when leaving the OR (a Joint commision requirement) that now we’ve done a complete turn in our practice. Also, having a sore nose/cheeks after wearing an N95 all day, day after day”

For some, the access to PPE was the major concern, combined with perceived lack of preparedness of the healthcare system to meet the challenges of a pandemic, and implied disregard for patient and staff safety:

> “At the very beginning of the pandemic I had less than zero support from my organization. I was ‘counseled’ by my division director because I was pushing my supervisors for support. … I was unable to get PPE for my nurse. I was unable to move my patient out of a double patient room. I was unable to get a negative pressure room for the patient. Moreover I was counseled for being concerned and raising my concerns because the patient did not meet ‘screening criteria’. … Two weeks later when we had a designated COVID unit, I was pulled there to work in close extended proximity with positive COVID patients and only given a surgical mask to wear for the full shift. The patients were unmasked. “

> “Having taken care of multiple patients without N95 masks immediately prior to them being diagnosed. The hospital does not notify staff when this happens and doesn’t seem to care that we are not protected. N95 masks are ridiculously hard to get a hold of.”

> “The lack of adequate PPE putting my friends and coworkers at risk due to poor planning. To me this is inexcuseable[sic].”

> “In the beginning, had NO N95 when it was clear that there were cases in the area and still had to see sick patients in the office. … Also initial lack of available testing. Was concerned about NH [nursing home] patients dying and staff getting sick, which happened and if we had more PPE and testing early on, would not have been as critical an issue”

A particularly difficult situations arose when it appeared that management refused to share the burden of risk with frontline workers who lacked PPE, actions perceived as cowardly or callous:

> “When we were first told we can only wear procedure masks. No goggles, and when we went in for a MATT the [leadership team] stood outside the door and just watched. Not one helped of them helped. … They let us go in and get contaminated but stayed in the hallway and watched.“

> “…RN grabbed an N95 and our Nurse manager told her she shouldn’t wast[e] PPE and didn’t need it. She said I’m protecting myself. The nurse manager said, you are protected this is what you signed up for when you became an RN.”

Although this concern is not necessarily specific to COVID-19 pandemic, some healthcare workers felt distressed by the degree to which PPE interfered with their ability to care of patients because of time it took to “properly place” it on when there were “so many patients and things to worry about”:

> “Most stressful was having to don and doff PPE each time before entering a patient room and trying to consolidate all the times necessary to enter/exit a room in the interest of preserving PPE/reducing exposure.”

### Concerns about health of oneself and others, not related to contracting COVID-19

Being unable to receive proper medical care due to infection control protocols in place proved very difficult for some:

> “I had a scheduled appointment with my [specialist] and it was very upsetting and stressful that they wanted to reschedule it due to the sole fact that I am … working with COVID patients.”

Some noted “lack of assistance getting access to mental health” as the causes of considerable distress, while others were affected by the increase in the mental health needs of their patients who exhibited “anxiety and emotional response to COVID-19” the led to “having to support them to ensure they do not become suicidal.”

For some, concern about health and stress of co-workers rose to the top of the most distressing experiences of the pandemic:

> “Seeing my co-workers stressed and suffer.”

> “… talking with employees who are stressed about this and trying to keep them rational…”

For a few, “unexpected death” of a family member during the pandemic overshadowed any other difficulties.

### Death of patients

For some healthcare workers, the sheer rate of mortality was overwhelming: “5 deaths in one shift.”

The circumstances of patients dying was among most difficult experiences, stressing the loneliness of these patients due to infection control protocols isolating them from their families:

> “Facetiming a mother whos daughter is dying and shes unable to be there because of restrictions”

> “I had to hold an Ipad over a dying man’s face while his wife and daughter screamed after we had coded him for over an hour and they had agreed to stop CPR for the last time. They were not able to be by his side so I had to let them have their final momentover face-time.”

> “so much death despite doing everything you can…sudden death…so much suffering for pts alone and for their families who can only facetime … to see the grief on the family”

> “Seeing nurses so defeated in having to facetime with families to help the loved ones say goodbye…. This is heartbreaking”

> “Watching patients struggling to breathe and watching the fear on their faces. Also families not being able to be present during this time.”

> “Holding the phone up to a barely responsive COVID positive dying elderly woman so her husband, unable to visit, could tell her he loved her and could say goodbye to her one last time-over the phone.”

> “Having to watch patients with covid die alone without family or a nurse or staff member at their bedside.”

The matters were especially difficult when the healthcare worker had a personal connection with the patient who died:

> “Having high risk pts that I have been following for years die from Covid.”

> “The patient was very uncomfortable, was suffering and could not breathe comfortably. No matter what I did, I could not help the patient to feel better. I stayed with her until she was admitted. Shortly after being admitted, the patient died. She had no past medical history, and she was one of my coworker’s family members.”

Some reported struggling with the sense of failure and futility of effort in trying to avert death of the COVID-19 patients, combined with belief that such patients could have received better care, including emotional support, even if they could not be saved:

> “working for two weeks , vigorously, only to have the patient die anyway after much struggle. Not having love ones updated by doctors, and having to tell them that they will not be able to see them even though I feel they are likely to die within days.”

> “Receiving patients who were dying from covid and I felt were suffering. Patients suffering from air hunger has been difficult to witness and even more difficult trying to make them comfortable in their dying hours. I feel haunted by some of the patients I worked with and am reminded often of their suffering.”

> “Very difficult to watch a patient suffocate slowly over weeks only to die despite our best efforts. No family is allowed to visit. Horribly sad.”

> “Holding hands with a small amount of patients as they take their last breath so that they won’t be alone as we continue to have visitor restrictions”

A recurring, experiences appeared to have been related to lapse in ethics of care due to lack of communication with families of patients:

> “That most people from nursing homes and the VA are dying and the DNR/AND is not honored. Families are not involved as much and do not understand what their loved one is being put through. People dying alone.”

Even some veterans of the field who “can handle almost anything” found they “cried everyday” when dealing with dying COVID-19 patients who “tend to be hopeless” and were denied usual level of care by infection control protocols:

> “Many of the patients do not recover totally or they die in the ICU. The patients have the commonality of severe respiratory distress even when intubated, sedated and on mechanical ventilation; this is difficult to observe. … the family members were extremely emotional on the phone, facetime or when the patient is coding. It was very difficult to participate in facetime and zoom with crying family members who could not hold a hand or touch their family member. I had to be the set of hands touching the patient while the family could not be present. “

Some choices that healthcare workers had to make, precipitated by infection control protocols, appear to be the stuff of ethical nightmares:

> “Patient likely going to die and having to limit which son will be able to remain.” “Having to Code elderly patients who had no hope of recovery.”

> “dying patient in the trauma bay and having to explain to a husband and daughter that they couldn’t be with their loved one in her final moments due to protocol”

### Perceived lapse (guilt over) in standards of patient care

Some healthcare workers were burdened with a sense of guilt over belief that they did not provide the usual level of care:

> “watching patients not be cared for in routine ways that would have been standard pre COVID has been particularly troublesome.”

> “felt a lot of guilt when patients that came in healthy got it and died.”

> “not being able to treat early covid patients properly due to no treatment.”

> “I felt as if I often did not have the time to give to my patients that they needed and deserved to be cared for properly.”

> “a traumatic intubation that I thought may have been too early to be necessary - before we learned that other methods might be more effective than intubation.”

> “I hated the idea of intubating clients because those clients all seemed to die. I felt like I was killing people.”

> “Video visits still very unwieldy and not easy to navigate for patients. Poor application.”

Some felt guilty when they believed that they let their teams down:

> “Injuring myself … . I felt like I was letting my colleagues down by not being at work and going through all the stressful changes with them.”

> “Taking a LOA due to [redacted: illness] was very difficult for me. I felt as though I was letting my co-workers and my community down because I was not present during the time when help was needed the most.”

Those who had work hours reduced reported that “not being able to provide nursing care in a pandemic has caused feelings of worthlessness.”

For some respondents, concerns about quality of patient care appear to have been aggravated by breakdown in teamwork and fairness in allocation of responsibilities:

> “Other people seem to be avoiding my ICU covid patients yet I am accused of not wanting to see them by a collegue. Meanwhile, my collegues I know aren’t seeing their patients and we are doing them a disservice by allowing services and doctors to NOT see their patients without repercussions. Its literally the most morally impossible situations I have been in in in my career.”

### Social isolation

For some healthcare workers, the most difficult experiences of pandemic arose from the loss of usual social contacts. In many cases this was related to self-isolation for fear of spread of hospital-acquired infection to family members at high risk from the virus, especially when combined with recognized mental health difficulties:

> “Dealing with my personal depression and anxiety, self isolation, not being able to see my mother in her SNF [skilled nursing facility]”.

> “Having to move out of my home/relocate due to COVID and to protect high-risk family members from me bringing it home to them.”

> “Being separated from my family to keep them safe. Stayed in hotel after shifts at hospital. When last shift completed I stayed additional 10 days before coming home.”

Isolation from family and wider community made coping with the pandemic more challenging for some, precisely because family, community and friends were the usual sources of support:

> “not being able to visit with my family and attend church.”

> “… it was extremely stressful to me that I was not able to see my family and friends who I often go to for support. I felt scared and guilty about seeing them knowing I would spend hours and hours in COVID positive rooms and did not want to infect anyone if I had brought it home with me from work.”

For those who rely on co-workers for support and companionship, having to stay away from work for fear of spreading infection to vulnerable family members was among most difficult experience of the pandemic, identifying such circumstances as the cause of depression:

> “Staying home again after extended leave and being isolated from everyone has lead to depression - on top of the extreme fear of my … baby contracting the virus.”

Work from home likewise result in erosion of social support from co-workers, such that “isolation due to working at home” was reported as being among more difficult experiences.

In some instances, healthcare workers felt ostracized by the community due to fear of contracting the virus, combined with perceived empty gestures of support and lack of tangible effort to make it easier for healthcare workers to function:

> “I had to deal with friend, neighbors considering me a deadly weapon because I might be infected and a threat to them. Everybody was at home, I had to come to work, nobody cared. I actually found the clapping etc demeaning, not a sign of support. I wish I could go to the grocery store without standing in line, but nobody thought that would be helpful for those working. This was not a very nice experience.”

> “-People (including other nurses ) looking at me as if I am infected with COVID just because I work on a COVID floor. - Social Isolation”

Difficult situations arose when healthcare workers felt that their ability to communicate risk of the pandemic with their patients was superseded by misleading media coverage, resulting self-censorship, which is a form of social isolation:

> “The media has created mass histeria. It’s snowballing. It’s hard to do my job when families are constantly throwing cnn or google in my face! Everyone is soo gulliable to take what they say as true. They don’t do any fact checking. This histeria has foced me to socially distant or i get sociall shamed. It just adds to the anxiety …”

### Stressed colleagues

Give the importance of teamwork to successful provision of healthcare, perceived stress of co-workers proved to be among most difficult experiences for some:

> “talking with employees who are stressed about … and trying to keep them rational…”

> “Seeing my co-workers stressed and suffer.”

> “Staff breaking down and crying; managing direct staff fears”

> “Dealing with irrational fears from colleagues and ancillary hospital staff.”

For some, breakdown in teamwork manifested in multiple ways, anchored in worries about risk of infection and unequitable workloads, and aggravated by lack of administrative support:

> “Staff worried about PPE every shift. Upper management not supportive to staff. Physicians not going into patient’s rooms. Nurses taking on the roles of multiple jobs.”

> “Co workers of other disciplines, in fear of entering a Covid-19 suspected positive or positive patients room, asking me to assess or perform tasks that are their job. Physicians, out of fear of coming into contact with a suspected or confirmed positive Covid-19 patient, refusing to enter the patients room or assess the patient.”

Unsupportive colleagues were a source of hardship when the whole team was facing challenging situations:

> “We had multiple deaths each week and docs were more intense and demanding than usual causing me more stress to the point of crying.”

Several healthcare workers appeared find it difficult to deal with colleagues who appeared to not be bearing well under what was perceived as normal pressures of intensive care:

> “Mostly hearing co-workers ‘fishing for thank you’s’. … being overly dramatic…I wourk in the intensive care setting on a daily basis and this is not hard.”

It appears that difficulties related to perceived “irrational fears” were conflated with frustration about how the pandemic was portrayed in the media and politicized, how such matters were seen as shaped by forces beyond healthcare workers’ influence:

> “How media has blown this out of proportion. They have created the assumption that a positive is a death sentence. … The media has created mass h[y]steria.”

> “The over the top societa[l], political response to a pedestrian disease”.

### Aggression and anger of patients

While dealing with stressed colleagues was among most difficult experiences for some, encounters with agitated and aggressive patients and their families was among the most adverse experiences of the pandemic for others, conferring a sense of not being “unappreciated”:

> “Being yelled and screamed at by distraught, isolated suspected and confirmed Covid-19 patients because they did not feel I was moving fast enough to garb up and answer their call bell or provide them with something they needed.”

It appears that for some aggression of patients was closely tied to worries about their endangering themselves and the healthcare workers:

### Uncertainty

Uncertainty of the impact of the virus on one’s patients appears to have intermingled with sense that patients are not receiving the best possible care:

> “Dealing with babies, and not know how it affects them. Not knowing if they get vertical transmission from their mothers. Not sure if they are positive with no symptoms in an open bay unit. Not sure if there are long term side effects of this to the newborn population… .”

> “I understand this is a terrible virus, I have struggled with the world being shut down for a virus with a 94%-98% cure rate. We didn’t shut the world down for far more lethal viruses. It has caused a ridiculous amount of stress isolating the people who need socialization the most. The elderly and the mental ill.”

> “Every day feeling like I am failing my patients because we don’t know enough, don’t have adequate plans in place, are not allowed to test in our outpatient office so we can’t give people good information or the care they desire.”

Difficulty ascribed to the uncertainties precipitated by the pandemic and response to it emerged in concerns for health of oneself and quality of patient care. However, it was not limited those areas but was more widespread, such as not knowing what the next day on the frontline of the pandemic will bring, a sense of apprehension:

> “Actually preparing myself for work. And knowing the stress that will ensue.”

> “Feeling that I will encounter situations that I am not confident in my ability to manage.”

> “That the only thing consistent about work is the inconsistency. You never know what you’re walking into and everything changes every day. And you never have answers.”

Lack of understanding of the new pathogen made it challenging for those in leadership positions to feel confidence in adequacy of their response:

> “As the director the biggest stress I had was the unknown about the virus. What kept me up at night was am I doing everything to keep my staff and patients safe and healthy.”

The uncertainty about what holds true about the pandemic made it difficult for some to train staff, when official policies contradicted person’s understanding of the evidence, making it difficult to “coach … staff on policies/initiatives that [one does] not personally agree with.”

Some healthcare workers reported that “managing staff anxiety, fear” was particularly stressful in light of “knowledge deficits”.

The constant changes, “at a daily and sometime hourly basis,” in the understanding of the pandemic at the leadership level precipitated stressful changes in procedures at the bedside:

> “One of most stressful situations I’ve had to deal with during the covid-19 pandemic was navigating the initial change in bedside care and policies.”

> “management and policy changes nearly daily that are directly impacting the health of staff, patients and peers. Blatantly idiotic decision making that flips daily but MUST be adhered to in the window of it’s lifespan. (do NOT wear an N95! next day…you MUST wear an N95…next day…you will NOT be fit tested…next day…you MUST be fit tested….next day…you do NOT need an N95 bc the CDC says so.)”

One respondent shared an experience of overcoming difficulties that arose from the stresses of chaotic initial period of the pandemic, by suggesting that one gained equanimity through continued exposure to the new conditions:

> “The unknown at the beginning of this was the most stressful to me because we didn’t know what we were exposing ourselves to or how to test people and if we had been exposed or not. I feel like I grew out of that by continuing to work … “.

For some, the diversity of views of the pandemic by the public and patients placed healthcare workers in the crossfire of the competing narratives, such that the community’s uncertainty had an adverse spill-over effect on the frontline healthcare workers:

> “interactions with people extremely paranoid about contracting the virus and interactions with people who don’t think the virus is real. The constant back and forth is emotionally exhausting.”

### Shortcoming in transparency at work

Some of the most difficult experiences related to “lack of communication”, “no answers, lack of answers, answers only to the individual asking”, including “not receiving information that the government and patients thinking [healthcare workers] had access to testing both for the virus and antibodies before [healthcare workers] did” and “no support from manager regarding how to find information or what to tell patients”. One healthcare worker summed this up as:

> “You never know what you’re walking into and everything changes every day. And you never have answers.”

When practice guidelines appeared to make no sense and work-related requests were not perceived as having been dealt with rationally and respectfully, distressing situations arose:

> “… being told I did not need to wear PPE. After push back, getting the required PPE and being questioned why I needed them.”

> “Having taken care of multiple patients without N95 masks immediately prior to them being diagnosed. The hospital does not notify staff when this happens and doesn’t seem to care that we are not protected.”

For some there was an overall sense of “the disorganization, poor communication, and frequent disrespect by [leadership/management]” which included perception of lack of concern for well-being of healthcare workers:

> “leadership ignoring staff input … Being told we are ’heroes’ while being treated like second class citizens.”

> “Absolutely horrendous communication from … central management with multiple conflicting communications and policies and NO input as to the formulation and implementation of these policies. It seems that a few people are hoarding the decision making process.”

> “When we were first told we can only wear procedure masks. No goggles, and when we went in for a MATT the [management, names redacted] stood outside the door and just watched. Not one helped of them helped. ”

It was suggested that “an organized explanation, and a respectful conversation is always best even in a ‘crisis’ situation”.

### Lack of administrative support

When frontline works appeared to be overlooked in recognition by the management despite taking risks and speaking up in an attempt to remedy lapses in practice, this proved “stressful”:

> “we were constantly exposed and not recognized at all. … The bullies … keep being bullies. Coming to work gives me anxiety and nothing is done. They have all been reported several times and they still have their jobs.”

Worse still, “being disciplined for being frustrated with the increased work loaf [sic]” was named among most difficult experiences of the pandemic, leading some to sense that they are “unable” to perform their duties both safely and well.

Some healthcare workers struggled with the experience summed up as “support from immediate supervisors has been lacking”, but other point out to specific lack of support in reducing the uncertainty and improving patient care, namely “no support from manager regarding how to find information or what to tell patients.”

Perceived lack of empathy and “respectful conversation” with administration was led to “unnecessary frustration”:

> “the disorganization, poor communication, and frequent disrespect by my director and the command center … if there was anxiety or questions asked by us, it was often a disrespectful response like, ‘you are lucky you have a job’, etc… no support or empathy if an employee felt unsafe with certain tasks given.”

> “Absolutely horrendous communication … with multiple conflicting communications and policies and NO input as to the formulation and implementation of these policies. It seems that a few people are hoarding the decision making process.”

When there was perception that rules were not fair, applicable to all, frustration arose due to:

> “Interacting with leadership that is not following set guidelines … - and being ridiculed for wanting to follow set guidelines.

“Lack of empathy and understanding” from leadership appears to be recurring theme in situations that caused hardships to healthcare workers, for example “when daycare closed and I had to figure out a schedule to still be able to work while my husband, who is also essential, was still able to work and our child was still cared for every day.” One of the expressions of this lack of empathy appears to have been seen in reports that

> “system is more worried about their bottom line than they are about patient safety and it shows: they’re cancelling all flex time and PTO for anyone not working on a COVID unit.”

Another expression of perceived lack of empathy is expressed in the experience of “lack of assistance getting access to mental health”.

### Childcare

Loss of childcare and the resulting need to balance professional duties as an “essential” worker with family responsibilities was among most difficult experiences of the pandemic reported by those with children living at home:

> “Working full time … while the schools have been closed for the last two months, my [children] are at home …”

This was especially difficult when person was the only available caregiver, such as due to illness of the other spouse and/or alternative caregivers being at a heightened risk from COVID-19:

> “My [spouse] was gravely ill with COVID and I had to care for both him and my children AND then return to work to care for my patients.”

> “My [spouse] has an immunodeficiency. I worry I’m going to bring home Covid … and that [they] will die. We have a … 2 years old… . His daycare is closed. It’s not a great idea to bring outside people (like a babysitter) into our home to to watch him. We can’t use grandparents because if I get them sick with covid, they are at risk of dying. My [spouse] and I both work. It’s been nearly impossible to work and care for our son. “

The issues with childcare were aggravated when management was not seen as supportive and understanding:

> “Lack of empathy and understanding from my boss when daycare closed and I had to figure out a schedule to still be able to work while my [spouse], who is also essential, was still able to work and our child was still cared for every day.”

### What has been the event that has most reinforced your pride in your professional behaviour?

Frequencies and names of the coded themes of the events since the start of the pandemic that reinforced pride in the profession are captured in **Table 2**. They also indicate our perceived association of these themes with autonomy-supportive factors experienced by healthcare workers. The nuances and meaning of the coded themes are illustrated blow using the respondents’ own words.

### Courage, composure under pressure

Given the anxieties and uncertainty of early days of the pandemic, it is not surprising that individuals who felt that they conquered their fears and remained on the job were proud of the fact:

> “… I haven’t allowed COVID to overtake my life with fear/anxiety”, “just dealing what comes next and driving on despite the deaths”, “not freaking out when learning i was possibly exposed”,

> “I gained the strength to come to work every day, even when I was frustrated and scared”,

> “How nurses have stepped up with courage and grace during this unprecedented time to provide compassionate, competent care to those ill with SARS-COV-2”.

> “I have tried to remain calm and practice positive mental health exercises”.

Give how taxing the work on the frontline of the pandemic proved to be, some were proud not only of the fact they continued to work but that they could say: “I still love the work I do”.

The pride in overcoming personal fears was related to putting the needs of patients above personal risks and hardships:

> “I’m afraid that I’m going to get COVID (and/ or bring it home to my family). But I have shown up to work every day and continued providing clinical care. It’s taken a lot of courage. But I do it.”

> “Showing up at work everyday, odd shifts despite the hardships of felling unprepared without all of the answers and constraints on family life (daycare closed, worried about my family, not seeing friends and neighbors).”

> “Going to work everyday knowing the risks, especially being respiratory compromised myself.”

For some respondents it appears that their pride in steadfastness of healthcare workers with intertwined with pride in effective teamwork:

> “The resolve of the … doctors and nursing staff … . There is a collegial feeling of purpose that is palpable but hard to articulate.”

> “Despite being burnt out and working short, our team has continued to show such support and strength. It lifted me up to continue to show up everyday during this trying time.”

> “The working staff sticking together and taking care of call and working weekends to cover those who were furloughed and for all of them keeping level heads.”

> “The staff that pull together, remain positive, and do whatever needs to be done to care for our patients while maintaining professionalism and a sense of humor.”

Some experiences that instilled pride combined the sense of being up to the challenge of the pandemic as the team, while receiving little support from the leadership of the healthcare system:

> “The way that nurses and doctors rose to the task at hand to fight the spread of Covid. We educated ourselves every chance we could get about the latest treatments. No one complained, we just did our job. We received no support or encouragement from our administration and they were never visible during this crisis.”

> “We succeeded in spite of administration. Not because of it.”

This pride in autonomous professional success is summarized succinctly as pride due to “relying On myself”.

Some noted that they were proud not in just how well they worked but by their professional growth:

> “My confidence has grown throughout this whole experience, knowing that I was able to be thrown into this crazy situation … and I was able to adjust and adapt to the constant daily changes all while being on a fully COVID floor for the entire time.”

### Volunteering for hazardous tasks and altruism

One clear example of courage under duress was expressing in volunteering for tasks perceived to be hazardous, an understandable source of personal pride:

> “I was an initial volunteer to be placed on a list of people who will strictly care for known Covid positive patients. I did this so that my co-workers who were too nervous to be in Covid rooms didn’t have to be.”

> “I will see covid patients to keep another provider, at higher risk that I, from seeing them”.

Some healthcare workers were proud of doing what they thought was right for their patients and colleagues, despite personal hardships and risks:

> “Knowing that I would do anything to make sure staff and coleagues have everything they need to remain safe”

> “Showing up at work everyday, odd shifts despite the hardships of felling unprepared without all of the answers and constraints on family life (daycare closed, worried about my family, not seeing friends and neighbors)”

> “Being able to be there for the community and treat patients while risking my own self and family’s health.”

This did not necessarily relate to volunteering for hazardous duties but taking extra effort to do what one thought was right, even when there was perceived lack of support from the administration for such acts:

> “When we were scolded by management for using the 3 cent bouffant caps in the covid rooms, I went home and cried. Then I made 60 reusable cotton (washable) caps that I donated to my peers. I makes me proud to see my caps on my peers 10 weeks later.”

> “Ultimately I had to use community resources and also my own money to buy [N95 masks and face shields] from to community also, but i’m proud I kept my staff safe. This was both very stressful and proud moment.”

Altruism and courage displayed by others was a clear source of pride about the profession overall:

> “seeing an 80+ year-old family medicine physician walking out of a COVID patient room in a PAPR. He could have not been there given his age, but he did not stay away. Inspiring.”

> “Everyone did what they had to do during the early aids epidemic before we knew what was going on some docs refused to treat patients we had none of that this time.”

> “To see all the ‘front line’ healthcare providers step right up without, any hesitation to personal risk, to provide care to our patients has been inspiring. People’s willingness to help work extra, take on new responsibilities and cover for those self-quarantined without any additional compensation during stressful times, reinforces my trust and belief in the wonderful colleagues I work with.”

### Quality of patient care, positive outcomes

Among most common experiences that reinforced professional pride among healthcare workers was the quality of care they provided, exemplified by positive outcomes:

> “the few who have been the most sick with covid and survived”.

> “Seeing many patients recover and get home safe.”

Beyond survival, specific aspects of patient care that were highlighted among those that instilled pride included educating and counseling patients to ease their fears:

> “successfully educating patients about what we currently know and easing their fear and anxiety”.

> “I do stay professional and not engage in argumentative conversations. I educate when appropriate and say little when that is appropriate. I realize all are afraid.”

> “Assisting patients through a very stressful health crisis. It is assuring to be able to walk patients through the process and see them come out well on the other end. Being able to offer a resource and support to patients in a confusing time.”

> “Having a Covid patient that was very ill and wanted to leave … and working with him through his anxieties and fears and supporting his fears and having him agreeable to stay for treatment and care.”

> “Listening to patients and their families’ needs in order to better serve them. Specifically when a fear of theirs can be alleviated.”

> “Being able to continue to care for patients and help them not feel abandoned by hcw due to fear of contracting illness”.

Pride in compassionate “high quality of empathetic, therapeutic care” was abundantly evident, especially needed due to isolation of patients precipitated by infection control measures:

> “being able to make these isolated pts smile, even if its just briefly”

> “The honor or caring for the patients during the tough days. Holding a hand, talking about their family and how they missed the ones they love.”

> “an elderly couple, married many years, both positive with COVID19, brought them together in same room, put their beds together so the husband could be with his wife at the end of her life”.

> “Helping a husband come in and see his very ill wife and seeing her react with a smile to him.”

> “Since I work in the nicu; doing everything I can to give parents footprints, photos, the camera”.

Some shared that they were proud of being able to help others provide the high-quality care:

> “That my former students at the bedside are setting examples of high quality, compassionate, and safe patient care”.

Success in infection control while providing high quality of care of infected patients was a clear source of pride:

> “Our office has seen greater volumes of patients who were having problems than others in the specialty and yet we are apparently the only office in the specialty that did not have a single employee covid case.”

> “Knowing that I have come to work through this all and have not contracted Covid.”

> “All of the people that have left alive after being infected and the fact that I have not gotten it and my family is all healthy and clean.”

Examples of assertiveness and successful advocacy on behalf of the patients were a source of pride for some, resonating with themes of pride in autonomous action, showing initiative:

> “I am proud of my ability to advocate for suffering patients to get physician orders for oxygen therapies and morphine drips.”

> “ultimately prevailing with clinical logic with administration to advocate for N95 masks and face shields … . Ultimately I had to use community resources and also my own money to buy some from to community also, but i’m proud I kept my staff safe. This was both very stressful and proud moment. also fighting to overturn the stupid … rule … . It does enhance bitterness at admin that they rule out policy before medical input. but we prevailed in the end.”

A related source of pride is how healthcare workers adapted to caring for patients, growing in both confidence and skill required to practice their profession under the pandemic’s constraints; there was a sense that this is what the profession was meant to do, a reinforced notion of fulfilling of one’s duty when it mattered most despite personal risks:

> “We were the first COVID unit. Watching the nurses go from being uncertain, scared to totally embracing it and doing an excellent job caring for these patients. Putting patients first. This is our calling, it’s why we are nurses.”

> “I love taking care of Covid patients b/c they are the most grateful and appreciative of all the patients. They need human contact and therapeutic touch, many do not give it to them b/c they are scared to and we’re told to have as little contact as possible. I give it to them anyway. I’m protected by my PPE. I’m a nurse patient care is why I went into nursing and this will not stop my practice.”

> “Fighting a disease that we know so little about and seemed to be everywhere and with numbers that were exponentially increasing all around us was very daunting. Working under the fear and anxiety that I would somehow expose my family was so incredibly stressful. But despite this, early on I had resolved to persevere and lead my team through this pandemic. I knew many people were counting on me to be a voice of calm, reason and guidance. After all, this is why I became a physician. To not run away in the face of an unknown infectious agent but towards it.”

Some shared that their pride rested on having provided quality care despite lack of recognition and support, simply due to sense of professional standards that the person set for themselves:

> “That even if no one gives a rip about what I did … . And I will continue to give the best care to my patients that I can, and work for them, and take pride in the job I do, to make our family medicine group into the best one around -- for the patient. It hurts that we were never recognized for the hard work we did, but we didn’t do it for the recognition anyway, but for our patients.”

> “I am very proud to be a part of my small community hospital, while administration, the WHO, CDC all left us with little to nothing in the way of policies and appropriate PPE, almost every one of my colleagues continue to come in each day and roll with the punches to take care of other human beings. It is stressful and we all feel like we are expendable.”

For many of the respondents, pride arose simply from doing their work despite pandemic:

> “I am still providing the same quality of care that I did before the pandemic and that is what I am proud of.”

### Comforting dying patients

Despite death of patients being named among most difficult experiences by healthcare workers, the manner they handled death of patients was a source of pride for many:

> “I sat with a covid patient who was put on hospice, held his hand, talked to him, kept him company and kept him comfortable while he was actively dying while his primary nurse was unavailable”.

> “Being able to be present for my patients during their final hours and having chance to call family members to speak with the patient before their passing has reinforced my pride in my professional behavior. In my opinion, that is one of the greatest things that we can do as a nurse - it shows that we are empathetic, compassionate, and caring. It takes a strong person.”

> “Being there for my patients. Talking to them even though they may not hear me. None of My patients died alone. I was there for each of them.”

The common element in these experiences that reinforced professional pride was the compassion and companionship offered to those who died.

### Innovation and technological sophistication

Some were clearly proud of their own inventiveness in overcoming challenges posed by the infection control protocols, reporting that they were proud of:

> “coming up with out of the box ideas … to facilitate better communication with patient and caregiver.”

> “… willing to try therapies that others feel maybe with unknow consequence but potential for significant benefit”.

The fact that developing new way of providing care necessitated initiative due to (perceived) lack of organizational support was a source of pride for some:

> “Working without any instruction on COVID patient care from hospital administration”.

### Community outreach, beyond clinical duties

Some respondents were proud of the fact that they contributed to management of the pandemic outside of their immediate clinical duties:

> “trying to be the middle ground, voice of reason and making sure I have researched facts I provide to friends/family/patients.”

> “development of worldwide information on COVID-19 that is in line with my beliefs … .”

> “WHEN A NURSE STOOD UP TO A PROTESTER PROTESTING THE RIGHT TO OPEN UP BUSINESSES.”

### Adaptability and flexibility

Healthcare workers reported to be proud of how they and their colleagues adapted to the challenges of the pandemic, reporting this to be seen as important quality care and effective teamwork:

> “Returned to the bedside and easily re-trained with success. I had a renewed sense of personal pride having done so.”

> “I have not only adapted to this rapid change but have overcome it. I am still providing the same quality of care that I did before the pandemic and that is what I am proud of.”

> “Everyone’s resourcefulness and support in coming up with ways to continue to take care of pts in ways that keep exposure to a minimum.”

For some, adaptability that they were proud of appeared to be facilitated by effective teamwork:

> “I have been very encouraged by the overwhelming support my colleagues have provided me during this time. Despite the changes, we all seem to adapt as best as we can and help each other throughout the days and weeks.”

> “Teamwork, sacrifice, and agility of each member of our staff to re-organize and implement provision of care while keeping patients and staff safe as possible.”

### Revealing leadership qualities

Some we proud of how they led their teams:

> “I took it upon myself to guide and take care of my team during time where office manager lacked decision making and / or communication with staff” (even when there was a chance to relinquish such a responsibility)

> “holding the staff together in this time, as they are very anxious …”

> “My ability to stay strong and be a calm leader in this crisis for my coworkers”, “I had resolved to persevere and lead my team through this pandemic. I knew many people were counting on me to be a voice of calm, reason and guidance.”

Some found effective leadership displayed by others a source of pride for the profession as a whole:

> “… the course shown by some nursing, physician and several other staffs.”

Those in the leadership positions appreciated being recognized when they proved effective:

> “Having the employees at the end of our conversations thank me for the help, when they say I feel a little better now”.

### Teamwork

Teamwork appears to be one of the most consistently and commonly reported sources of pride in how healthcare workers comported themselves during the pandemic, despite fears and uncertainties. This is exemplified by a report “that nurses and other health care providers, even when scared without having information, still came to work and did their job for their patients.” The sense of *esprit de corps* and focus on the mission to serve patients despite personal risk is clearly articulated in this narrative:

The pride in teamwork arose not only because it was reported to lead to best possible care for the patients but because it entailed peer support: “Happy with support of co-workers at coming together to work in a pandemic.”

### Support from community

For some, pride stemmed from recognition of healthcare workers by the wider community, which included not only expressions of thanks but also material support:

> “The outpouring of community support, donations, thank yous, and the complete strangers that were kind enough to make scrub hats for us.”

> “All of the outpouring from the community at large. Even the commercials on television provide me with pride.”

> “We as a team received a lot of support from local businesses supplying lunches and words of encouragement. It helped the long shifts filled with uncertainty and stress a little easier to get through.”

> “Seeing the parade put on by cops/fire trucks and ambulances. Was surprised by how much that moved me”.

Recognition of difficulties faced by healthcare workers by media (and thus community at large) appeared to be an important part of instilling a sense of pride:

> “I appreciate the truth about the lack of crucial medical supplies and PPE nurses and medical personnel are forced to work without everyday; putting our own health at risk, being reported in the news. In addition, I appreciate the news reporting how medical staff and nurses are treated when we try to ourselves safe and healthy by advocating for ourselves and speaking up for what is happening that is not right.”

Expressions of gratitude and concern that had a personal touch appeared to be appreciated:

> “We received cards from children, churches and even families as being hero’s at the bedside. A previous patient from last fall family member called the unit, in February, before the influx of patients, to ask if the nurses are “OK” because he was worried about us. I did not know this person, but I became emotional when a stranger had concerns related to staff members future endeavors and COVID.”

> “Having a man at a Wawa sincerely thank me for helping to keep people safe”.

Expressions of support and gratitude from family members who thereby assumed person risk of infection were likewise a source of pride:

> “My family supporting me in continuing my job as a nurse, despite being away from home, while friends were telling me to get out of nursing due to COVID.”

> “The community and my family/friends support was truly amazing. I felt appreciated for the work I was doing”.

### Gratitude from patients

The sense of professional pride was reinforced when patients expressed gratitude for care that they received, with special emphasis on quality of communication, including managing worries:

> “The gratitude expressed by patients, especially when I’m able to help ease their fears and anxiety. Best part is being able to discharge someone who was positive for COVID and has since recovered.”

> “compliments from my patients, when they tell me they appreciate me listening to them and taking the time needed to take great care of them”.

It appears that gratitude from some patients and their families was related to both quality of care and recognition of personal risks that healthcare workers endured in order to care for the patients:

> “A woman with [condition that is not COVID-19 redacted] said to me with tears in her eyes after her surgery, “Thank you for being here for me. With all that is going on, I was so worried that there wouldn’t be anyone to take care of me.”

### Recognition from administrators and leadership of healthcare systems

Healthcare workers who believed to have been appreciated by their employers and leaders within their organizations tended to report this as one of the experiences that reinforced pride in their professions:

> “Weekly uplifting emails from management, positive posts on social media. Immediate responses to problems faced with concrete solutions.”

> “Receiving a quality award for exceptional performance”

When organizations did not come across as divided between healthcare workers and administration, there was a reason to report this as a proud achievement:

> “The love, support and spirit of my nurses, first responders, and hospital administration.”

For some healthcare workers, the pride in their professions was reinforced by acts of advocacy by those in a position to do so:

> “… our director fought to keep us open to care for patients who still needed immediate treatment. “

> “my union’s persistence”

### Not proud

Some respondents elected to tells us why there were not proud of their profession, even though we did not inquire about this, indicating that there is a strong sentiment of failure and shame among some respondents. Some of this relates to regret of not having done enough:

> “Retrospectively, I am honestly not proud of my professional behavior. I should have tried harder to get the necessary resources … in the early days of the infection. I should not have let the counseling deter me for advocating for myself and my colleagues.”

The sense of not being able to advocate for the best handling of the pandemic appears to be a common them among those who appear to feel less than proud about their profession’s and their personal contribution during pandemic, with the source of frustration aimed at government and hospital leadership:

> “Government is causing the numbers to be inflated because they are paying hospitals for every patient diagnosised with CoVID, even more if they are intubated or die as the diagnosis. It’s wrong.”

Others expressed dismay at how healthcare systems reacted, conveying a sense of futility and apathy in light of a contradiction between known best practices and demands of administration/leadership:

> “If anything, everything I’ve seen since the start of COVID has made me believe that professionalism is gone from this hospital. I try to keep it together for my patients, but who cares about PPE regulations …We know this is wrong, but our administration tells us to do it anyway- so why try?”

Some responses appear to capture a sense of exhaustion, burn-out:

> “I dont have any feelings any more to my profession. I just take one day at a time. Having feelings is not much use.”

It appears that for some respondents pride in the profession depended on action of external forces and how they were treated rather than intrinsic values or achievements:

> “Pride is out the window. We have been treated increasingly worse as this rolls on. Most staff are barely hanging on and several have quit.”

It would appear that for some the value of recognition within organizations and faith in rational actions in the best interest of patients outweighs positive impact of support from community at large:

> “Pride is dead in this profession. Media calls us heroes while leadership treats us like an expendable commodity. … MANY patients and staff exposed due to incompetent decisions … .”

